# Methods for joint modelling of longitudinal omics data and time-to-event outcomes: Applications to lysophosphatidylcholines in connection to aging and mortality in the Long Life Family Study

**DOI:** 10.1101/2024.07.29.24311176

**Authors:** Konstantin G. Arbeev, Olivia Bagley, Svetlana V. Ukraintseva, Alexander Kulminski, Eric Stallard, Michaela Schwaiger-Haber, Gary J. Patti, Yian Gu, Anatoliy I. Yashin, Michael A. Province

**Affiliations:** Biodemography of Aging Research Unit, Social Science Research Institute, Duke University, Durham, North Carolina 27708, USA; Department of Chemistry, Washington University in St. Louis, St. Louis, Missouri 63130, United States; Department of Medicine, Washington University in St. Louis, St. Louis, Missouri 63130, United States; Center for Metabolomics and Isotope Tracing at Washington University in St. Louis, St. Louis, Missouri 63130, United States; Taub Institute for Research on Alzheimer’s Disease and the Aging Brain, Vagelos College of Physicians and Surgeons, Columbia University, New York, New York 10032, USA; G.H. Sergievsky Center, Vagelos College of Physicians and Surgeons, Columbia University, New York, New York 10032, USA; Department of Neurology, Vagelos College of Physicians and Surgeons, Columbia University, and the New York Presbyterian Hospital, New York, New York 10032, USA; Department of Epidemiology, Mailman School of Public Health, Columbia University, New York, New York 10032, USA; Division of Statistical Genomics, Department of Genetics, Washington University School of Medicine, St. Louis, Missouri 63110, USA

**Keywords:** lysophosphatidylcholines, aging, mortality, longitudinal omics, repeated measurements

## Abstract

Studying relationships between longitudinal changes in omics variables and risks of events requires specific methodologies for joint analyses of longitudinal and time-to-event outcomes. We applied two such approaches (joint models [JM], stochastic process models [SPM]) to longitudinal metabolomics data from the Long Life Family Study focusing on understudied associations of longitudinal changes in lysophosphatidylcholines (LPC) with mortality and aging-related outcomes (23 LPC species, 5,790 measurements of each in 4,011 participants, 1,431 of whom died during follow-up). JM analyses found that higher levels of the majority of LPC species were associated with lower mortality risks, with the largest effect size observed for LPC 15:0/0:0 (hazard ratio: 0.715, 95% CI (0.649, 0.788)). SPM applications to LPC 15:0/0:0 revealed how the association found in JM reflects underlying aging-related processes: decline in robustness to deviations from optimal LPC levels, better ability of males’ organisms to return to equilibrium LPC levels (which are higher in females), and increasing gaps between the optimum and equilibrium levels leading to increased mortality risks with age. Our results support LPC as a biomarker of aging and related decline in robustness/resilience, and call for further exploration of factors underlying age-dynamics of LPC in relation to mortality and diseases.

## INTRODUCTION

Contemporary longitudinal studies on humans started collecting repeated measurements of various omics (e.g., metabolomics, proteomics) data for study participants. Availability of other types of information on the participants such as follow-up data on mortality and onset of diseases, genetic markers, questionnaires, repeated measures of health-related biomarkers, etc., provides extensive opportunities to study complex relations of individual age-trajectories of omics variables with risks of diseases and mortality, in connection to various genetic and non-genetic factors. However, this abundance of information and opportunities comes along with many methodological challenges related to analyses of such massive data. One particular complication deals with an inherent complexity of analyzing trajectories of health-related variables (repeated measurements of omics variables provide a good example of such) in relation to time-to-event outcomes. The Cox model with time-dependent covariates [1] is the conventional approach traditionally used for joint analyses of time-to-event data and repeated measurements of covariates. However, it is well known that it has certain limitations: ignoring measurement errors or biological variation of covariates and using their observed “raw” values as time-dependent covariates in the Cox model may lead to biased estimates and incorrect inferences [2-4], especially when covariates are measured at sparse examinations or with a long time interval before an outcome event. This applies to analyzing repeated omics measurements in relation to time-to-event outcomes as well. Even though relevant biostatistical methods, known as joint models (JM) [4, 5], have found broad applications in different research areas, their use in analyses of longitudinally measured omics data is still limited to a few small-sample proteomics studies [6-8].

One particular class of models for joint analyses of longitudinal and time-to-event outcomes, the stochastic process model (SPM), has been developed in the biodemographic literature based on the mathematical foundations laid out in [9-11]. Recent developments in SPM methodology merged the statistical rigor of the general approach with biological soundness of specific assumptions built into its structure [12-15] (see [16] for non-technical introduction to SPM), This brings the biological content to the model structure that made such models particularly appealing for research on aging. The main advantage of using SPM for research on aging is that it allows disentangling a general association between the longitudinal and time-to-event outcomes that can be found using JM into several components representing specific aging-related characteristics embedded in the model. This allows researchers not only to evaluate mortality or incidence rates but also to estimate age-related changes in the mechanism of homeostatic regulation of biological variables, the age-related decline in adaptive capacity and stress resistance, effects of allostatic adaptation, and allostatic load. This provides a more detailed perspective on the impact of the longitudinal dynamics of the respective variables on the risk of the modelled events in the context of aging. Despite broad applications of SPM to different outcomes and biomarkers (see, e.g., [13, 17-23]), to date, there were no applications of SPM to analyses of longitudinal omics measurements in relation to time-to-event outcomes.

In this paper, we fill these gaps and apply both JM and SPM to longitudinal measurements of metabolomics in more than 4,000 participants of the Long Life Family Study (LLFS) [24]. To illustrate applications of the approaches, we focus on a particular class of lipid metabolites, lysophosphatidylcholines (LPC), that have been actively discussed in the literature in relation to cardiovascular, infectious, and neurodegenerative diseases, and tested as potential early markers of Alzheimer’s disease and accelerated aging [25-31]. Overall, the literature suggests (see, e.g., the recent review [32]) that the reported LPC findings are somewhat contradictory because most of the recent studies, in contrast to older ones, found lower LPC levels to be associated with unfavorable outcomes such as mortality. In addition, longitudinal dynamics of LPC in relation to mortality and aging-related outcomes remains understudied. Here we aimed to test general associations of different LPC species with total (all-cause) mortality in the LLFS using JM and to investigate how such general associations can be decomposed into relations of the mortality risk with different aging-related characteristics (such as robustness, resilience, age-specific norms, and allostatic trajectories [16]), and whether such relationships/characteristics differ by sex.

## RESULTS

### Applications of the basic JM

Table 2 shows results of applications of the basic JM (Eqs. 1–2) [4, 5, 33] to measurements of LPC species and mortality data in the LLFS. The table presents the values of the association parameter (*α* in Eq. 1) for the respective metabolites in the survival sub-model, along with corresponding hazard ratios (for a unit increase in log-transformed and standardized metabolites) and their 95% confidence intervals (CI) estimated from the JM adjusted for the covariates indicated in **Joint models: Specific versions used in applications**. For the majority of LPC species (19 in the total sample, 16 in females, and 17 in males), the estimates of the association parameter *a* are negative and CI for respective HR do not contain one. This means that the increase in the levels of these metabolites reduces mortality risk. The strongest association in the total sample in terms of the point estimate was observed for LPC 15:0/0:0 (*α* = -0.335, HR = 0.715). This metabolite was selected for additional analyses illustrating different specifications of JM and more detailed investigation of its association with different aging-related characteristics embedded in the structure of SPM; see below. Supplementary Figure S1 displays diagnostic plots assessing the goodness-of-fit and assumptions of JM in applications to LPC 15:0/0:0. Figure S1 (a) shows random behavior of standardized marginal residuals around zero (with 95.65% of values lying within the (−1.96, 1.96) interval) validating the assumptions for the within-subjects covariance structure in the longitudinal part of JM. The Cox-Snell residuals plot (Figure S1 (b)) also shows the overall good fit of the survival sub-model of JM. Supplementary Table S2 provides estimates of other parameters in the longitudinal and survival sub-models from the basic JM applied to LPC 15:0/0:0. For comparison, Supplementary Tables S3-S4 show respective estimates in the Cox model with LPC used as a time-dependent covariate and all other covariates as in the survival part of JM. One can see from these tables that, even though the direction of effects and their significance are preserved in most cases, the association parameters are biased towards smaller magnitudes in the Cox model as expected from prior research [3, 34].

**Table 1:** Characteristics of the Long Life Family Study metabolomics subsample used in the analyses.

| Characteristics | Field Center |  |  |  | Total Sample |
| --- | --- | --- | --- | --- | --- |
|  | BU | NY | PT | DK |  |
| Number of families | 235 | 248 | 216 | 78 | 573 |
| Number of participants at any visit | 1,128 | 721 | 1,115 | 1,047 | 4,011 |
| Number of participants at visit 1 | 1,070 | 636 | 1,055 | 895 | 3,656 |
| Number of participants at visit 2 | 570 | 361 | 534 | 669 | 2,134 |
| Number (%) of deceased participants | 382 (33.9%) | 291 (40.4%) | 417 (37.4%) | 341 (32.6%) | 1,431 (35.7%) |
| Follow-up period (years) (mean ± SD [range]) | 10.3 ± 4.4<br>[0.28, 17.00] | 9.8 ± 4.1<br>[0.52, 17.00] | 10.3 ± 4.4<br>[0.29, 17.00] | 11.5 ± 4.8<br>[0.55, 17.00] | 10.5 ± 4.5<br>[0.28, 17.00] |
| Age at baseline (mean ± SD [range]) | 70.0 ± 15.8<br>[32, 110] | 74.2 ± 15.8<br>[24, 108] | 71.5 ± 15.9<br>[36, 104] | 69.5 ± 14.4<br>[38, 104] | 71.0 ± 15.6<br>[24, 110] |
| Whites (%) | 99.6% | 98.8% | 99.6% | 99.5% | 99.4% |
| Females (%) | 56.5% | 54.4% | 55.9% | 55.9% | 55.8% |
| Low educated participants (below high school) (%) | 6.0% | 7.8% | 7.1% | 29.3% | 12.7% |
| Smokers (smoked >100 cigarettes in lifetime) (%) | 41.0% | 45.5% | 36.2% | 48.9% | 42.6% |
| *APOE* ε4 allele carriers (%) | 15.3% | 19.1% | 17.8% | 26.3% | 19.6% |
| Medication use: angina (%) | 34.1% | 33.0% | 34.0% | 26.5% | 31.9% |
| Medication use: anti-diabetic (%) | 6.8% | 6.9% | 9.4% | 6.1% | 7.4% |
| Medication use: anti-hypertensive (%) | 52.7% | 56.0% | 56.9% | 47.8% | 53.2% |
| Medication use: lipid-lowering (%) | 35.5% | 45.9% | 40.4% | 24.0% | 35.8% |

**Notes:** a) Number of missing data: race – 20, education – 10, smoking – 18, *APOE* – 257, angina medications – 273, anti-diabetic drugs – 273, anti-hypertensive drugs – 273, lipid-lowering drugs – 273, other variables listed in the table have no missing values; b) The numbers shown in “Number of deceased participants” and “Follow-up period” correspond to the LLFS data release used in this paper (see **Data**). Abbreviations: BU – Boston, DK – Denmark, NY – New York, PT – Pittsburgh, SD – standard deviation.

**Table 2:** Results of applications of joint models to measurements of LPC species and mortality data in the LLFS: Estimates of the association parameter for the metabolite in the survival sub-model.

| Metabolite | Total |  | Females |  | Males |  |
| --- | --- | --- | --- | --- | --- | --- |
|  | Alpha | HR (95% CI) | Alpha | HR (95% CI) | Alpha | HR (95% CI) |
| LPC 0:0/16:0 | -0.222 | **0.801 (0.718,0.894)** | -0.183 | **0.833 (0.717,0.967)** | -0.185 | **0.831 (0.740,0.934)** |
| LPC 0:0/16:1 | -0.019 | 0.981 (0.870,1.105) | -0.036 | 0.965 (0.816,1.141) | -0.028 | 0.972 (0.837,1.130) |
| LPC 0:0/18:0 | -0.038 | 0.963 (0.872,1.064) | -0.029 | 0.971 (0.862,1.093) | -0.082 | 0.921 (0.786,1.080) |
| LPC 0:0/18:1 | -0.150 | **0.861 (0.779,0.952)** | -0.109 | 0.897 (0.764,1.053) | -0.151 | **0.860 (0.770,0.960)** |
| LPC 0:0/18:2 | -0.233 | **0.792 (0.704,0.892)** | -0.153 | 0.858 (0.727,1.013) | -0.330 | **0.719 (0.603,0.856)** |
| LPC 0:0/20:3 | -0.164 | **0.849 (0.764,0.942)** | -0.160 | **0.852 (0.775,0.936)** | -0.161 | 0.851 (0.716,1.011) |
| LPC 0:0/20:4 | -0.192 | **0.825 (0.746,0.913)** | -0.250 | **0.779 (0.671,0.905)** | -0.131 | 0.877 (0.765,1.006) |
| LPC 0:0/22:6 | -0.237 | **0.789 (0.711,0.876)** | -0.301 | **0.740 (0.642,0.854)** | -0.173 | **0.841 (0.726,0.974)** |
| LPC 14:0/0:0 | -0.236 | **0.790 (0.681,0.917)** | -0.185 | **0.831 (0.712,0.969)** | -0.212 | **0.809 (0.674,0.972)** |
| LPC 15:0/0:0 | -0.335 | **0.715 (0.649,0.788)** | -0.300 | **0.741 (0.650,0.844)** | -0.267 | **0.766 (0.667,0.879)** |
| LPC 16:0/0:0 | -0.241 | **0.786 (0.694,0.890)** | -0.190 | **0.827 (0.719,0.952)** | -0.164 | **0.849 (0.763,0.945)** |
| LPC 16:1/0:0 | -0.045 | 0.956 (0.854,1.071) | -0.049 | 0.952 (0.807,1.124) | -0.054 | 0.947 (0.812,1.104) |
| LPC 17:0/0:0 | -0.206 | **0.814 (0.729,0.908)** | -0.182 | **0.834 (0.731,0.951)** | -0.292 | **0.747 (0.610,0.914)** |
| LPC 18:0/0:0 | 0.031 | 1.031 (0.935,1.137) | 0.026 | 1.026 (0.912,1.154) | 0.000 | 1.000 (0.852,1.174) |
| LPC 18:1/0:0 | -0.209 | **0.811 (0.734,0.895)** | -0.177 | **0.838 (0.729,0.963)** | -0.171 | **0.843 (0.759,0.936)** |
| LPC 18:2/0:0 | -0.241 | **0.786 (0.711,0.868)** | -0.158 | **0.854 (0.742,0.983)** | -0.188 | **0.829 (0.750,0.917)** |
| LPC 18:3/0:0 | -0.163 | **0.850 (0.724,0.999)** | -0.030 | 0.970 (0.779,1.207) | -0.341 | **0.711 (0.553,0.915)** |
| LPC 20:2/0:0 | -0.226 | **0.798 (0.715,0.889)** | -0.160 | **0.852 (0.738,0.984)** | -0.280 | **0.756 (0.642,0.890)** |
| LPC 20:3/0:0 | -0.277 | **0.758 (0.680,0.845)** | -0.259 | **0.772 (0.671,0.890)** | -0.324 | **0.723 (0.589,0.887)** |
| LPC 20:4/0:0 | -0.212 | **0.809 (0.728,0.900)** | -0.221 | **0.802 (0.693,0.928)** | -0.202 | **0.817 (0.702,0.950)** |
| LPC 20:5/0:0 | -0.334 | **0.716 (0.657,0.780)** | -0.354 | **0.702 (0.602,0.819)** | -0.246 | **0.782 (0.676,0.905)** |
| LPC 22:5/0:0 | -0.237 | **0.789 (0.717,0.868)** | -0.265 | **0.767 (0.666,0.882)** | -0.201 | **0.818 (0.727,0.920)** |
| LPC 22:6/0:0 | -0.269 | **0.764 (0.692,0.844)** | -0.315 | **0.730 (0.632,0.842)** | -0.233 | **0.792 (0.690,0.909)** |

**Notes:** **Alpha** – estimates of the association parameter for the longitudinal variable (metabolite) in the survival sub-model of the joint model (parameter $\alpha$ in Eq. 1); **HR** – hazard ratios (for a unit increase in log-transformed and standardized metabolite values) computed from the association parameters; **95% CI** – 95% confidence intervals for HRs (highlighted in **bold** are cases where confidence intervals do not contain one). The joint models were estimated using R-package *JM*.

### Applications of JM with random intercept and slope

The results of applications of the general JM described in the previous section established the associations of the LPC species with mortality risk. Here we use different specifications of JM (Eqs. 3–5) with individual intercepts and slopes of (log-transformed and standardized) LPC that provide a further look at the relationships between the age dynamics of LPC 15:0/0:0 and mortality risk. Table 3 presents the results of applications of the models with individual intercepts (“int”, Eq. 4) and individual intercepts and slopes (“intslope”, Eq. 5) of LPC 15:0/0:0 to the total sample. Supplementary Table S5 provides estimates for sex-specific samples. The values of LPC_randomint in the tables show the values of the regression parameter (*α*_0_) for the random intercept (*b*_*0i*_) in the hazard rate (Eqs. 4–5). The negative estimates of the parameter indicate that larger baseline levels of LPC 15:0/0:0 are associated with reduced mortality risks (after adjusting for relevant covariates, see in the Survival part of the tables). This was observed in both models (“int” and “intslope”) and in total and sex-specific analyses. Similarly, there are negative estimates for LPC_randomslope, which is the value of the regression parameter (*α*_l_) for the random slope (*b*_l*i*_) in the hazard rate (Eq. 5), but the association was not significant (see also Note under Supplementary Table S5 about CI for males in “intslope”). The negative estimates of this parameter mean that faster changes in LPC 15:0/0:0 with age might be associated with reduced mortality risks (in the model adjusting for the covariates indicated in the Survival part of the tables). In the next section, we will further decompose the associations of LPC 15:0/0:0 and mortality risk considering different aging-related components embedded in the structure of SPM that provide additional details in the context of the aging process.

**Table 3:**
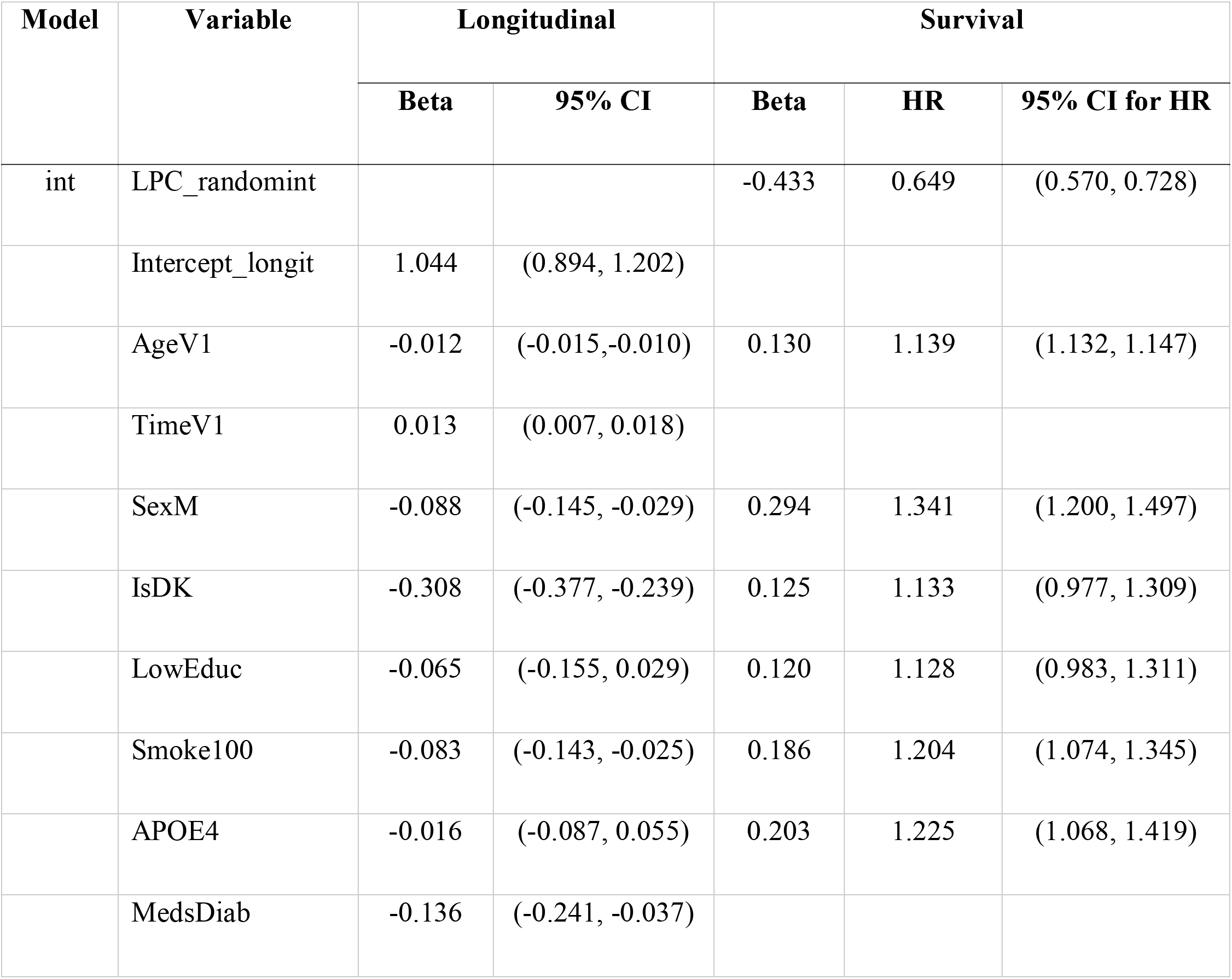
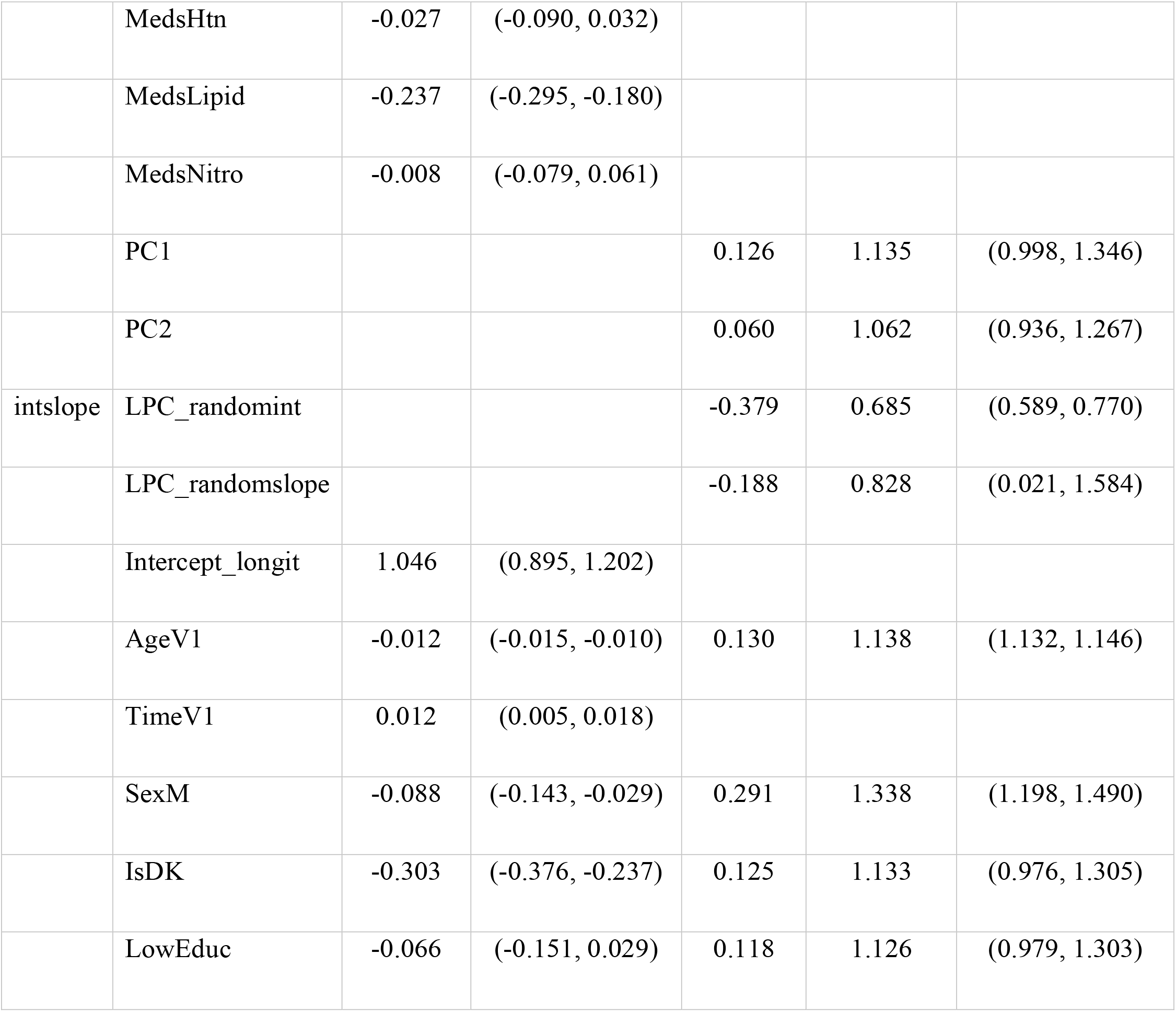
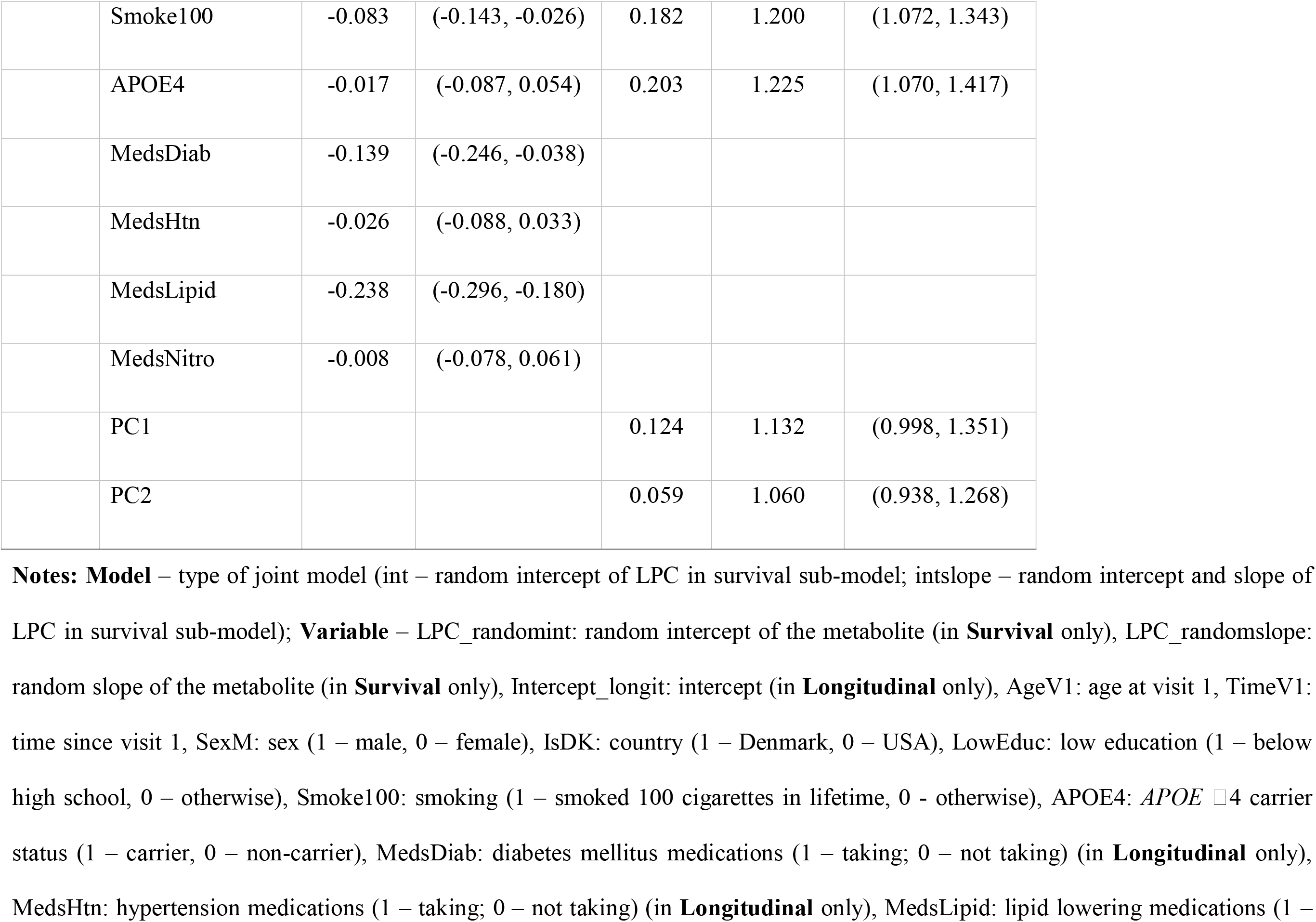
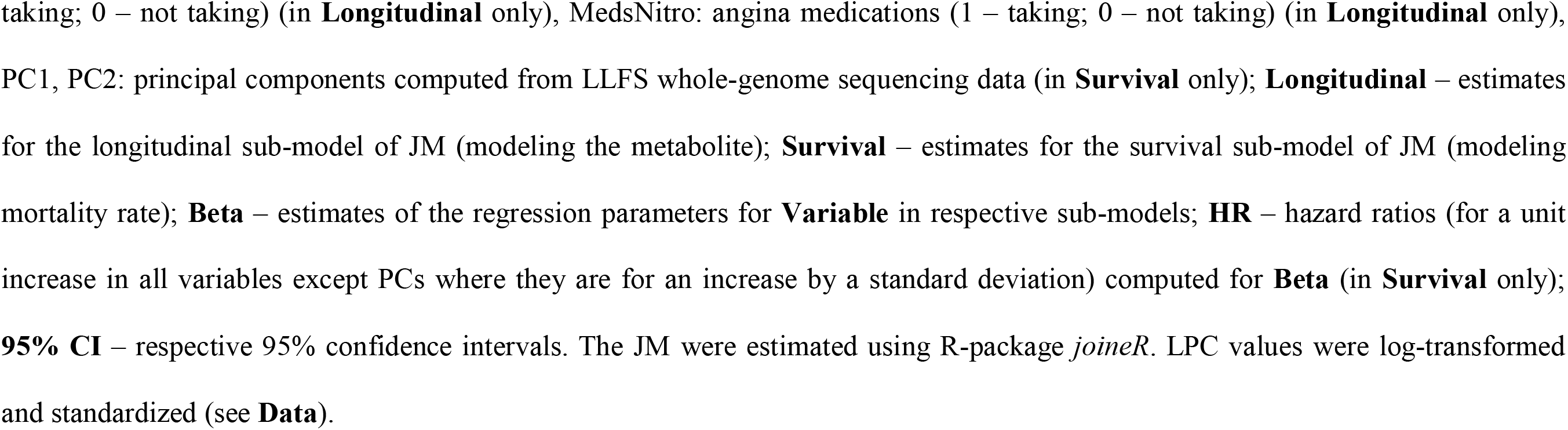
Results of applications of joint models with random intercept and slope of LPC 15:0/0:0 in mortality rate in the LLFS.

### Applications of SPM

Table 4 presents and interprets the results of testing various null hypotheses (H0’s) in applications of SPM to measurements of (log-transformed and standardized) LPC 15:0/0:0 and mortality in the LLFS metabolomics sample. Supplementary Table S6 contains estimates of parameters in the main (unrestricted) model and in different restricted models fitted for testing those hypotheses. Figure 1 displays each estimated model’s components evaluated for females and males along with *p*-values corresponding to different H0’s. Supplementary Figure S2 provides 3D plots illustrating the results of SPM. Figure S2a presents a 3D plot of the mortality rate as a function of age and LPC 15:0/0:0. It shows an increase in the mortality rate for lower levels of LPC 15:0/0:0 confirming findings in JM. Figure S2c displays the quadratic part in the mortality rate, that is, the difference between the total mortality rate and the baseline mortality rate (which does not depend on LPC 15:0/0:0, shown in Figure S2b). This additional term is larger for older individuals with lower levels of LPC 15:0/0:0. However, on the relative scale (i.e., the ratio of the total mortality rate and the baseline mortality rate), the largest ratio is at younger ages (where mortality is low) and for lower levels of LPC 15:0/0:0 (Figure S2d).

**Table 4:** Results of applications of stochastic process models to measurements of LPC 15:0/0:0 and mortality data in the LLFS: Results of testing different null hypotheses on patterns of model’s components.

| Null hypothesis<br>(H0) | LR statistics | DF | P-value | Interpretation of findings |
| --- | --- | --- | --- | --- |
| $Q(t, c) = 0$ | 100.382 | 3 | <0.0001 | Rejection of this H0 justifies further applications of SPM to LPC 15:0/0:0 and mortality (otherwise, there would be no quadratic term in the hazard rate [Eq. 7] so that the dynamics of LPC 15:0/0:0 would be unrelated to the mortality risk). |
| $Q(t, c) = Q(c)$ | 12.590 | 1 | 0.0004 | U-shape of the mortality risk as a function of LPC 15:0/0:0 narrows with age (Figure 1a). That is, as individuals grow older, they become more vulnerable to deviations of LPC 15:0/0:0 from the optimal trajectory $f_0(t, c)$ because the same deviation of LPC 15:0/0:0 from $f_0(t, c)$ results in a larger increase in the mortality risk at older ages compared to younger ages. |
| $Q(t, c) = Q(t)$ | 1.346 | 1 | 0.25 | U-shape of the hazard rate (as a function of LPC 15:0/0:0) does not depend on sex, i.e., there is no sex difference in robustness related to trajectories of LPC 15:0/0:0. |
| $a(t, c) = a(c)$ | 0.000 | 1 | 1.00 | Feedback coefficient ($a(t, c)$ in [Eq. 6]) is age-independent, i.e., there is no age-related decline in adaptive capacity (resilience) related to trajectories of LPC 15:0/0:0. |
| $a(t, c) = a(t)$ | 25.318 | 1 | <0.0001 | Males have better adaptive capacity (resilience) to deviations of LPC 15:0/0:0 from the equilibrium trajectory $f_1(t, c)$ compared to females (Figure 1b). Larger absolute values of $a(t, c)$ in [Eq. 6] for males mean a faster return of the trajectory of LPC 15:0/0:0 to $f_1(t, c)$ in case of deviations from it (i.e., better adaptation or resilience), compared to females. |
| $b(t, c) = b(t)$ | 24.026 | 1 | <0.0001 | Females have higher variability of LPC 15:0/0:0 (Figure 1c). |
| $f_1(t, c) = f_1(c)$ | 195.890 | 1 | <0.0001 | Equilibrium trajectories of LPC 15:0/0:0 decline with age (Figure 1d). |
| $f_1(t, c) = f_1(t)$ | 13.446 | 1 | 0.0002 | Females have higher levels of LPC 15:0/0:0 than males (Figure 1d). |
| $f_0(t, c) = f_0(c)$ | 3.898 | 1 | 0.048 | The optimal trajectory of LPC 15:0/0:0 that minimizes the mortality risk increases with age (Figure 1e). |
| $f_0(t, c) = f_0(t)$ | 0.678 | 1 | 0.41 | The optimal trajectory of LPC 15:0/0:0 does not depend on sex, i.e., LPC levels minimizing mortality risks coincide for females and males. |
| $f_1(t, c) = f_0(t, c)$ | 79.528 | 3 | <0.0001 | There is a gap between the equilibrium and optimal trajectories of LPC 15:0/0:0 (Figure 1f). |
| $f_1(t, c) = f_1(c)$ and<br>$f_0(t, c) = f_0(c)$ | 199.788 | 2 | <0.0001 | The gap between the equilibrium and optimal trajectories of LPC 15:0/0:0 increases with age (Figure 1f). Such deviating patterns lead to an increased mortality risk compared to the situation when the metabolite follows the optimal trajectory $f_0(t, c)$. |

**Notes:** LR – likelihood ratio, DF – degrees of freedom. LPC values were log-transformed and standardized (see **Data**).

**Figure 1:**
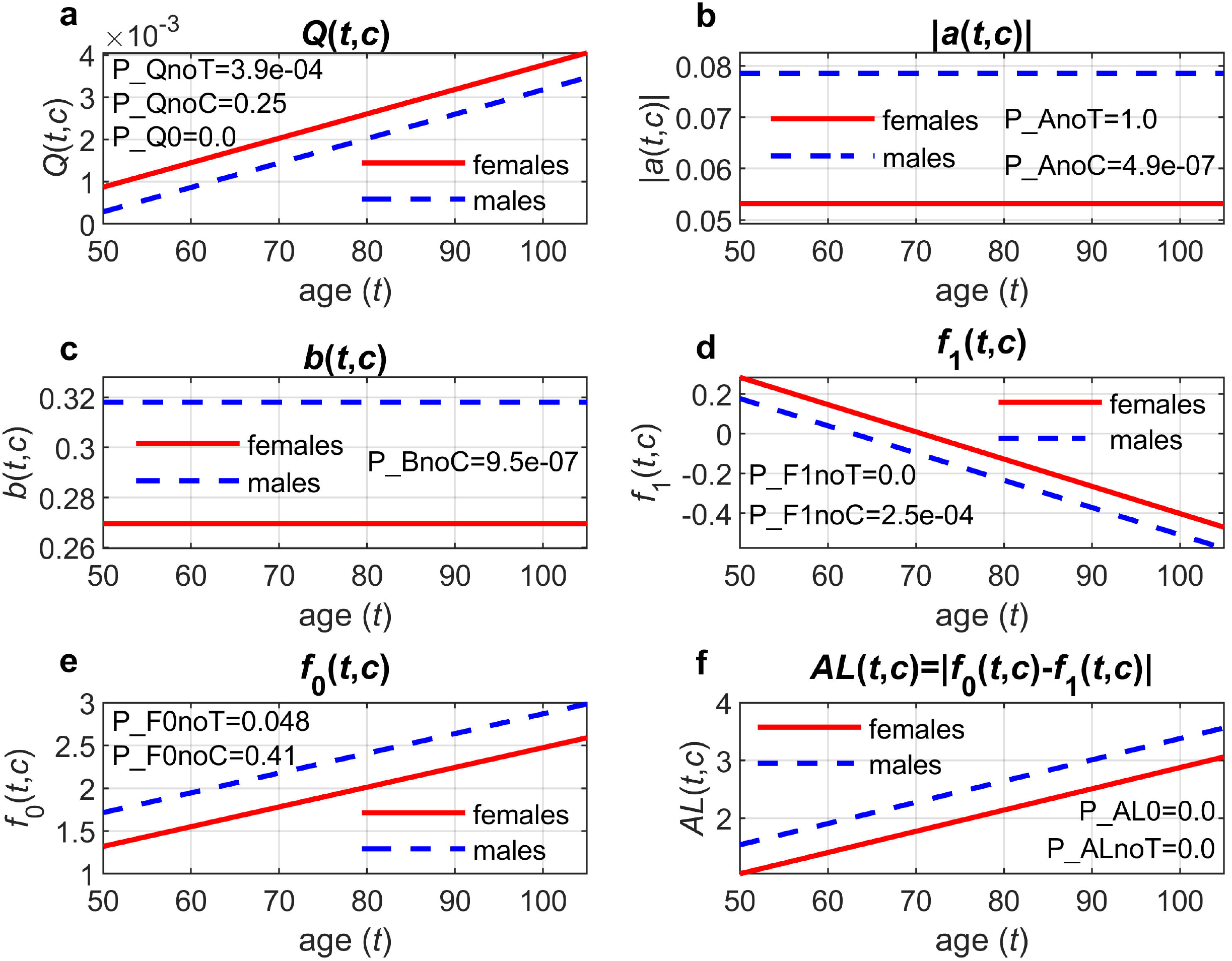
Applications of stochastic process models to measurements of LPC 15:0/0:0 and mortality data in the LLFS: Estimates of different components of the model. a) quadratic hazard term (*Q*(*t, c*)); b) adaptive capacity (|*a*(*t, c*)|); c) diffusion coefficient (*b*(*t, c*)); d) mean allostatic trajectory (*f*_*1*_ (*t, c*)); e) optimal trajectory (*f*_0_(*t, c*)); f) measure of allostatic load (*AL*(*t, c*) *=* |*f*_0_(*t, c*) *-f*_1_(*t, c*)|); *p*-values shown on the graphs are for different null hypotheses (H0): H0: *Q*(*t, c*) *= Q*(*c*) (P_QnoT); H0: *Q*(*t, c*) *= Q*(*t*) (P_QnoC); H0: *Q*(*t, c*) *=0* (P_Q0); H0: *a*(*t, c*) *= a*(*c*) (P_AnoT); H0: *a*(*t, c*) *= a*(*t*) (P_AnoC); H0: *b*(*t, c*) *= b*(*t*) (P_BnoC); H0: *f*_1_(*t, c*) *= f*_1_(*c*) (P_F1noT); H0: *f*_1_(*t, c*) *= f*_1_(*t*) (P_F1noC); H0: *f*_0_(*t, c*) *= f*_0_(*c*) (P_F0noT); H0: *f*_0_(*t, c*) *= f*_0_(*t*) (P_F0noC); H0: *f*_1_(*t, c*) *= f*_0_(*t, c*), i.e., *AL*(*t, c*) *=0* (P_AL0); H0: *f*_1_(*t, c*) *= f*_1_(*c*) and *f*_0_(*t, c*) *= f*_0_(*c*), i.e., *AL*(*t, c*) *= AL*(*c*) (P_ALnoT). LPC values were log-transformed and standardized (see **Data**).

In brief, we found that:

1. U-shape of mortality as a function of LPC 15:0/0:0 narrows with age so that older individuals become more vulnerable to deviations of LPC 15:0/0:0 concentrations from the trajectory of its optimal values.
2. Males have better resilience to deviations of LPC 15:0/0:0 from the equilibrium (“mean allostatic”) trajectory compared to females.
3. Females have higher variability of LPC 15:0/0:0.
4. Equilibrium trajectories of LPC 15:0/0:0 decline with age.
5. Females have higher equilibrium levels of LPC 15:0/0:0 than males.
6. The optimal values of LPC 15:0/0:0 that minimize the mortality risk increase with age.
7. There is a gap between the optimal and equilibrium trajectories and this gap increases with age.

### Sensitivity analyses

Supplementary Table S7 presents estimates of JM using the familial bootstrap approach [35]. While there were a few cases where 95% CI for HR in the main calculations (Table 2) did not contain 1.0 but the HR range in the familial bootstrap included 1.0 (highlighted in yellow in Supplementary Table S7) and one opposite case (highlighted in grey in Supplementary Table S7), in most cases the sensitivity analysis confirmed the results shown in Table 2. In particular, LPC 15:0/0:0, which was selected for downstream analyses, showed strong associations with mortality (e.g., HR=0.743 range: [0.585, 0.849] in the combined females + males analyses). Supplementary Table S8 presents estimates of SPM in the familial bootstrap analyses. The results are similar to the original computations (cf. Supplementary Table S6), in both the direction and the magnitude of effects. The resulting patterns of the model components are similar to those shown in Figure 1 (data not shown).

## DISCUSSION

This work is the first application of two approaches dealing with joint modelling of longitudinal and time-to-event outcomes (JM and SPM) to a large-scale metabolomics study, which collected repeated measurements of metabolomics for more than 4,000 participants. These approaches allow performing statistically rigorous analyses of repeated measurements of omics data jointly with time-to-event outcomes avoiding common pitfalls of the traditional tools that ignore biological variability/measurement errors in longitudinal outcomes and informative missingness arising because of attrition due to mortality (which is a common situation due to the nature of the outcomes studied in research on aging) [2, 3, 16]. The basic JM [4] which we used in our applications allows establishing general associations of longitudinal omics variables with time-to-event outcomes by including the “true” values (i.e., the difference between the observed value and the error term, see Eq. 2) of the variable in the hazard rate and computing respective hazard ratios. The JM version capturing associations between the longitudinal and time-to-event outcomes by a latent Gaussian process [36-38] provides different specifications of associations that include individual intercepts and slopes of omics variables in the hazard rate (Eqs. 3-5). These are the ones from the random part of the longitudinal sub-model of JM, i.e., they represent individual characteristics after adjustment for covariates (in the fixed part of the longitudinal sub-model) and the error term. Such models expand analyses by the basic JM and quantify the relations between these individual characteristics of omics trajectories with time-to-event outcomes, e.g., computing hazard ratios for a unit increase in an individual slope. The SPM digs deeper into the relations between the longitudinal dynamics of omics variables and time-to-event outcomes decomposing the associations observed in JM into several components representing relevant aging-related characteristics. Such characteristics include biological/physiological norms (“sweet spots” [39-41]), allostatic trajectories and allostatic load, as well as age-related decline in adaptive response to deviations from allostatic (equilibrium) trajectories and age-related increase in vulnerability to deviations from the norms, which represent decline in biological/physiological robustness and resilience considered key manifestation of aging [42]. SPM analyses thus can shed more light on relations between age trajectories of omics variables and time-to-event outcomes in the context of aging.

Our applications of JM to data on repeated measurements of different LPC species and mortality in the LLFS found that, for the majority of LPCs, larger levels were associated with reduced mortality risk (or, equivalently, lower levels were associated with increased mortality risk), in the total sample as well as in separate analyses in females and males. This confirms recent results that reported associations of lower LPC levels with unfavorable health outcomes including mortality, see, e.g., reviews in [25, 32]. For example, in the study of patients with sepsis [43], the non-survival group had significantly lower levels of LPCs 16:0, 17:0, and 18:0 compared to the survival group. In the study involving acute-on-chronic liver failure patients [44], those who died had lower LPC levels than survivors. Decreased LPC levels were significantly associated with increased mortality in bacterial community-acquired pneumonia patients [45]. Reduced LPC levels were associated with poor prognosis (including mortality) in individuals with acute liver failure [46]. All these prior publications reported findings in small samples from specific groups (patients with different diseases/conditions). To the best of our knowledge, our work is the first study that confirmed associations of LPC species and total (all-cause) mortality in a large longitudinal study with thousands of participants and repeated metabolomics measurements.

SPM applications illustrated how the observed associations between the LPC species (taking as an example the variant with the strongest association, LPC 15:0/0:0) and mortality found in JM reflect underlying aging-related characteristics that shape the observed age trajectories of LPC and their impact on mortality risk. In particular, we found that the U-shape of the mortality risk as a function of LPC 15:0/0:0 narrows with age reflecting aging-related decline in robustness to deviations of trajectories of LPC 15:0/0:0 from the optimal levels (that is, those minimizing the mortality risk at a given age). That is, the same magnitude of deviation at older ages leads to a larger increase in the risk than at younger ages. We also observed sex differences in the ability of an organism to return to the equilibrium (mean allostatic) levels of LPC 15:0/0:0: males revealed a better adaptive capacity so that in males, it takes less time for LPC 15:0/0:0 levels to return to equilibrium levels in case of deviations from them, compared to females. These estimated equilibrium levels differ by sex with females having higher (more favorable in terms of the mortality risk) levels than males. The equilibrium levels also decline with age whereas the optimum levels show an increasing pattern with age. As result, there is an increasing gap between the optimum and equilibrium levels, which leads to an increased mortality risk with age. One particular advantage of SPM is that it allows evaluating optimal levels or ranges [16] of longitudinal outcomes (e.g., LPC species as in our applications). Such levels/ranges derived from the model can take into account potential confounders and conceptualize the “optimum” as the levels minimizing the mortality risk (or risks of other events of interest, which, hypothetically, can differ). Such optimal levels do not necessarily coincide with average sex-specific levels for particular ages as our SPM analyses of LPC 15:0/0:0 illustrate. Thus, SPM applications can expand and complement the ongoing efforts to compute reference values of metabolites for different ages and sexes [47] and can provide additional information that can be used in clinical decision-making processes.

Our SPM results are in line with other studies exploring the LPC-aging nexus. Recently, it was found [31] that higher levels of LPC species were associated with slower biological aging (expressed by two DNA methylation-based metrics). In particular, the LPC species with 15 carbons showed the strongest (negative) association with the biological aging metrics in that study. Lower baseline concentrations and faster decline in levels of several LPC species were associated with faster decline in skeletal muscle mitochondrial function in longitudinal analyses [48]. The impaired mitochondrial oxidative capacity was previously found to be related to lower levels of several LPC species [49]. Older adults with dual decline in memory and speed showed the most extensive alterations (faster decline) in LPC metabolic profiles [50]. The anti-oxidative stress and anti-inflammatory responses have been suggested as potential biological mechanisms that can explain the observed associations of LPCs with slower biological aging [25, 31, 32]. However, as noted in the recent study [51], LPCs can exhibit opposite signatures, both anti-inflammatory and pro-inflammatory, so that their impact on health can be more ambiguous with potentially pleiotropic or competing roles that may depend on physiological context, comorbidities or other factors such as age. As our SPM applications indicate, sex can also be a significant factor contributing to various hidden aging-related characteristics underlying the LPC trajectories and their relations to mortality. Analyzing sex differences in LPCs (and phospholipids in general) in relation to aging in longitudinal cohort studies is of considerable interest and importance because of the paucity of such studies, and, in particular, considering inconsistencies regarding sex differences in LPC levels during aging observed in prior research [52]. Impacts of other factors on the observed relations between LPC trajectories and mortality can be explored using the tools in this paper including the genetic underpinnings of the relationships that can be evaluated using relevant tools [14, 53].

We acknowledge that our study has limitations. First, we used simple specifications for the models, e.g., linear functions of age for SPM components. While versatile and flexible, such specifications do not allow exploring more complex non-linear age patterns of respective characteristics. We are limited in our choice by the current availability of repeated measurements of metabolomics in LLFS (up to two per individual). Second, the LLFS is predominantly (>99%) a white sample. Therefore, our findings need confirmations in other studies collecting data for other race/ethnicity groups. Third, we performed analyses of a single metabolite in one-dimensional SPM. While multivariate JM and SPM are available [15, 54], their practical applications in analyses of samples similar in size to this study can be intractable. Relevant dimensionality reduction techniques (e.g., as in our prior works [34, 55]) can be used to mitigate this. Fourth, we used available tools developed for analyses of unrelated samples. Even though sensitivity analyses using the familial bootstrap confirmed the robustness of our results, development and validation of approaches handling relatedness among study participants can benefit future analyses of longitudinal omics data in family-based studies.

## MATERIALS AND METHODS

### Data

The Long Life Family Study (LLFS) [24] is a family-based, longitudinal study of healthy aging and longevity that enrolled participants at four field centers (three in the US: Boston, New *Y*ork, Pittsburgh, and one in Denmark). The LLFS recruited 4,953 individuals from two-generational families selected for exceptional familial longevity based on the Family Longevity Selection Score [56]. The first in-person evaluation (Visit 1) was done in 2006–2009. The second in-person visit (Visit 2) of surviving participants from Visit 1 and newly enrolled participants was completed in 2014–2017. Visit 3 started in 2020 and is ongoing. The participants provided information on socio-demographic indicators, past and current medical conditions, medication use, and physical and cognitive functioning [24]. Annual telephone follow-up was conducted to collect updates on participants’ vital and health status. All reported deaths were adjudicated by an Adjudication Committee [24]. We used the September 20, 2023 release of the phenotypic LLFS data, with the latest recorded follow-up date on June 29, 2023. Baseline ages were validated using dates of birth from official documents in the US [57] and through the civil registration system in Denmark. Ages at censoring for those alive at the end of the follow-up period were determined from dates of birth and the last follow-up. Ages at death/censoring and an indicator of death were used as time-to-event outcomes in our applications of SPM. In JM applications, time since the baseline was used as the time variable due to the specifics of the JM software used in the analyses.

We used batch 6 (released on October 25, 2023) of LLFS metabolomics data, which provides information on 188 lipid metabolites measured longitudinally in the LLFS participants at Visits 1 and 2. In total, the LLFS metabolomics sample contains 6,776 measurements of the metabolites (4,221 in Visit 1 and 2,555 in Visit 2). Plasma samples were first processed by using solid-phase extraction kits with both aqueous and organic solvents [58]. Extracted metabolites were then analyzed with liquid chromatography/mass spectrometry (LC/MS). To assess lipid metabolites, reversed-phase chromatography was used in combination with an Agilent 6545 quadrupole time-of-flight mass spectrometer at Washington University in St. Louis. A combination of different tools was used to remove background, annotate adducts, and identify compounds [59-61]. Missing values were imputed using the half-minimum approach (i.e., zeros were replaces by half of the minimum value) [62]. Profiling was performed in batches of approximately 90 samples. Batch correction was accomplished by using a random forest-based batch correction algorithm [63], which outperformed other approaches in the lipid metabolite data [58]. The metabolites were annotated by using standardized names from RefMet, version 07/2023 [64]. We used 23 available lysophosphatidylcholines (LPC) as the longitudinal outcomes in the analyses described below: LPC 0:0/16:0, LPC 0:0/16:1, LPC 0:0/18:0, LPC 0:0/18:1, LPC 0:0/18:2, LPC 0:0/20:3, LPC 0:0/20:4, LPC 0:0/22:6, LPC 14:0/0:0, LPC 15:0/0:0, LPC 16:0/0:0, LPC 16:1/0:0, LPC 17:0/0:0, LPC 18:0/0:0, LPC 18:1/0:0, LPC 18:2/0:0, LPC 18:3/0:0, LPC 20:2/0:0, LPC 20:3/0:0, LPC 20:4/0:0, LPC 20:5/0:0, LPC 22:5/0:0, LPC 22:6/0:0. Each metabolite was analyzed separately in one-dimensional models. Intensity values were (natural) log-transformed and standardized (to have a zero mean and a unit variance) before use in the models. The characteristics of the LLFS metabolomics sample are presented in Supplementary Table S1. Table 1 describes the analytic sample obtained after removal of records with missing information on covariates (see Notes under Table 1), which comprised 4,011 participants with 5,790 measurements of each metabolite (3,656 in Visit 1 and 2,134 in Visit 2, with 2,232 participants having one measurement and 1,779 participants with two measurements); 1,431 participants died during the follow-up period.

### Joint models: General specifications

The basic form of JM [4, 5] as implemented in the R-package *JM* [33] jointly estimates the parameters of the longitudinal trajectories of LPCs and mortality rates. The survival part of JM represents the mortality rate as a function of a metabolite and other covariates:

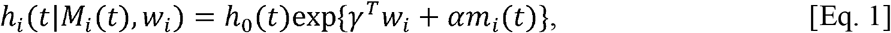

where *h*_*i*_ (*t*| ·) is the mortality rate for *i*^th^ individual at time point *t, m*_*i*_(t) is the “true” (i.e., unobserved) LPC level (see below in the text after Eq. 2) at time *t, h*_0_(·) is the baseline mortality rate, *w*_*i*_ is a vector of baseline covariates (see below), *γ* is a vector of respective regression coefficients, and *α* (a scalar) is the association parameter for the “true” LPC level (with respective hazard ratio, HR, for a unit increase in the “true” LPC level, computed in the traditional way as *HR* = *exp*(*α*)). A linear mixed effects model describes changes in the LPC levels as a function of time and other covariates:

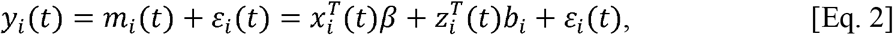

where *yi* (*t*) is the observed LPC level at time point *t* in the *i*^th^ individual, *x*_*i*_ (*t*) and *z*_*i*_ (*t*) are corresponding fixed and random effects, β and *b*_*i*_ are the respective vectors of parameters (that model population- and individual-level characteristics of LPC trajectories, respectively), and *ε*_*i*_(t) is the error term (independent of *b*_*i*_), normally distributed with zero mean and variance σ^2^ The difference between the observed value *y*_*i*_(*t*) and the error term *ε* (*t*), 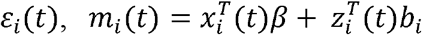, represents the “true” LPC level included in Eq. 1.

In addition to the general form of JM as in Eqs. 1–2, we used the JM versions where the association between the longitudinal and time-to-event outcomes is captured by a latent Gaussian process [36-38], as implemented in the R-package *joineR*, which allows different specifications of associations of individual dynamics of metabolites with the mortality rate. The general formula for the longitudinal part is as in Eq. 2, but the expression for the hazard rate differs from Eq. 1:

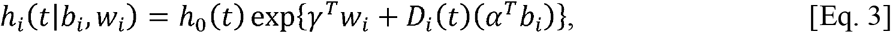

where *α* is a vector of association parameters corresponding to random effects *b*_*i*_ and *D*_*i*_(*t*) is the corresponding design matrix. We used two specifications of the JM: the intercept model (“int”) and the intercept and slope model (“intslope”). In the latter case, we used the option “*sepassoc=TRUE*” in function *joint* from the R-package *joineR*. In the “int” model,

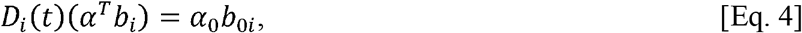

that is, the individual intercept of LPC (representing individual differences in baseline levels of LPC) enters the hazard rate, and in the “intslope” model,

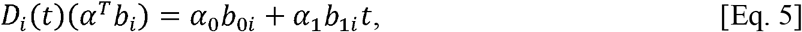

i.e., the individual intercepts and slopes of LPC (representing individual differences in both baseline levels of LPC and rates of change in LPC levels over time) are both tested for their association with the mortality rate.

### Joint models: Specific versions used in applications

In our applications of the basic form of JM [33], the longitudinal trajectories of different LPC species were modelled by a linear mixed effects model (Eq. 2) with linear random effects, i.e., random intercept and random slope. Time since the baseline visit was used as a time variable (as implemented in the R-package *JM*). Additional covariates were included in the fixed effects part of the longitudinal sub-model of JM: sex (1: male, 0: female), age at baseline visit, country (1: Denmark, 0: USA), education (1: below high school, 0: otherwise), smoking (smoked >100 cigarettes in lifetime: yes [1]/no [0]), medication use (anti-diabetic, lipid-lowering, anti-hypertensive, heart disease) (1: used, 0: not used), and APOE4 (1: carriers of apolipoprotein E [*APOE*] □4 allele; 0 – non-carriers of □4). The medications listed above include all available groups constructed by the LLFS investigators from original medications records using the Anatomical Therapeutic Chemical Classification System codes.

The time-to-event outcome (i.e., the mortality rate) was modeled as in Eq. 1, with the same covariates as in the longitudinal sub-model except medications (which are time-dependent). In addition, two genetic principal components (PCs) were included as covariates in the hazard rate (we tested models with different numbers of PCs and the results were similar; data not shown).

The baseline mortality rate *h*_0_(*t*) was modelled by a piecewise constant function. The pseudo-adaptive Gauss-Hermite quadrature rule [65] was used to approximate the required integrals in the estimation procedure.

In the specification of JM implemented in the R-package *joineR*, we used the same list of covariates in the longitudinal and survival sub-models. Unlike the *JM* package, the baseline hazard is represented semi-parametrically in *joineR*. Individual values of random intercepts and slopes were used in the expression of the hazard rate as shown in Eqs. 3–5, instead of the “true” level of the metabolite as in Eq. 1.

R version 4.3.1 was used to run the R-packages *JM* (version 1.5-2) and *joineR* (version 1.2.8) estimating respective models.

### Stochastic process models: General specifications

For SPM applications, we used a one-dimensional version with time-dependent components [15]. The dynamics of a repeatedly measured variable (*Y*(*t, c*), where *t* is age and *c* denotes covariates) is represented as a stochastic process with the following equation (in our applications, this equation models age trajectories of LPC):

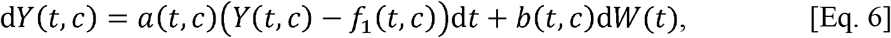

with initial condition *Y*(*t*_0_, *c*). Here *W*(*t*) is the stochastic (Wiener) process (assumed to be independent of *Y*(t_0_, c)) that defines random paths of *Y*(*t, c*), *b*(*t, c*) is the diffusion coefficient controlling the variability of *Y*(*t, c*), *f*_l_(*t, c*) is the long-term mean of the stochastic process, and *α* (*t, c*) is the negative feedback coefficient regulating how fast the trajectory of *Y*(*t, c*) returns to the mean *f*_l_(*t, c*) when it deviates from it. The SPM expresses the hazard rate (i.e., the mortality rate in our case) as a function of age (*t*), the vector of covariates (*c*) and the value of the longitudinal variable *Y*(*t, c*):

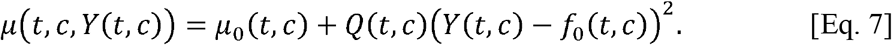

Here *μ*_0_(*t, c*) is the baseline hazard (i.e., mortality in our case) rate, *Q*(*t, c*) is the multiplier scaling the quadratic term of the hazard at different ages and values of covariates, and *f*_0_(*t, c*) represents the values of *Y*(*t, c*) (i.e., LPC) minimizing the risk (mortality) at age *t* and covariate values *c*.

The main characteristic feature of SPM is that Eqs. 6–7 embed several aging-related concepts (see more details in [16, 17]) thus facilitating more detailed analyses and interpretation of results in the context of aging, compared to analyses by JM: a) *homeostatic regulation*, which is a fundamental feature of a living organism; b) *allostasis and mean allostatic* (*“equilibrium”*) *levels* (*f*_l_(*t, c*)), featuring the effect of allostatic adaptation [66], i.e., the LPC levels forced by organism’s regulatory systems functioning at non-optimal levels; c) *adaptive capacity* (*α* (*t, c*)), modelling the rate of adaptive response (associated with *biological resilience* [17, 42, 67]) to any factors causing deviations of *Y*(*t, c*) from their dynamic equilibrium levels *f*_l_(*t, c*); d) *physiological or biological optimums* (*“sweet spots”* [39-41]) naturally represented by *f*_0_(*t, c*); e) *vulnerability component of stress resistance* (associated with *biological robustness* [67-70]) captured by the U-(J-)shape of the hazard and regulated by the multiplier *Q*(*t, c*) in the quadratic part of the hazard; f) *allostatic load* (*AL*) computed as *AL*(*t, c*) = |*f*_0_(*t, c*) *-f*_l_(*t, c*)| and representing the practical realization of the theoretical concept of AL suggested in the literature [66, 71-73] (the larger the value of this AL measure, the larger the price or load is for an organism in terms of an increased mortality risk compared to the best case scenario in which the trajectory of LPC follows the optimal function *f* _0_(*t, c*), i.e., when AL = 0).

### Stochastic process models: Specific parameterizations used in applications

For applications, we used the following specification of SPM: a) the Gompertz baseline hazard (represents a common pattern of mortality rate at adult and old ages): 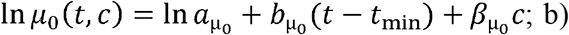 constant diffusion coefficient (based on our prior simulations showing the best accuracy of parameter estimates for models with constant *b*(.) [15]): *b*(*t, c*) = *σ*_1_ + *β*_W_*c*; and c) linear functions of age for other components (to estimate age trends in the respective components): *a*(*t, c*) = *a*_*Y*_ + *b*_*Y*_(*t - t*_min_)+ *β*_*Y*_*c*, where *a*_*Y*_ *< 0*, b_Y_ ≥20 and 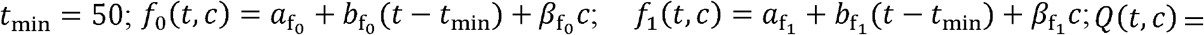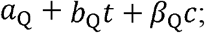 and 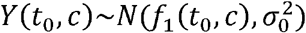 Based on our prior experience (dictated by technical complexities of the estimation algorithm), we included all covariates used in JM in *μ*_0_(*t, c*), whereas only one covariate (sex) was included in all other components.

In-house MATLAB codes (run in MATLAB version R2023a) implementing estimation algorithms with covariates in discrete-time approximations of SPM [18, 55] were used to estimate SPM parameters. Likelihood ratio tests were used to test several null hypotheses about the functional forms of each model’s components (i.e., to test whether they depend on age and on respective covariates). First, an “unrestricted” model (with the parameterization presented above) with no restrictions on parameters was estimated. Then, other models that contain one or more restrictions on parameters were estimated to test respective null hypotheses (H0’s):

1. H0: *Q*(*t, c*) =*0* (Qzero; interpretation: no quadratic term in the hazard);
2. H0: *Q*(*t, c*) = *Q*(*c*) (QnoT; the term in the quadratic hazard does not depend on age, i.e., robustness to deviations of LPC from the optimal trajectory does not depend on age);
3. H0: *Q*(*t, c*) = *Q*(*t*) (QnoC; the term in the quadratic hazard does not depend on sex, i.e., robustness to deviations of LPC from the optimal trajectory does not depend on sex);
4. H0: *a*(*t, c*) = *a*(*c*) (AnoT; the feedback coefficient does not depend on age, i.e., the adaptive capacity [resilience] is age-independent);
5. H0: *a*(*t, c*) = *a*(*t*) (AnoC; the feedback coefficient does not depend on sex, i.e., the adaptive capacity [resilience] is the same in females and males);
6. H0: *b*(*t, c*) = *b*(*t*) (BnoC; the diffusion coefficient is age-independent);
7. H0: *f*_1_(*t, c*) = *f*_1_(*c*) (F1noT; equilibrium LPC levels are the same for all ages);
8. H0: *f*_1_(*t, c*) = *f*_1_(*t*) (F1noC; equilibrium LPC levels do not differ by sex);
9. H0: *f*_0_(*t, c*) = *f*_0_(*c*) (F0noT; LPC “sweet spots” are the same for all ages);
10. H0: *f*_0_(*t, c*) = *f*_0_(*t*) (F0noC; optimal LPC levels minimizing mortality risks coincide for females and males);
11. H0: *f*_1_(*t, c*) = *f*_0_(*t, c*), i.e., *AL*(*t, c*) =*0* (ALzero, zero AL);
12. H0: *f*_1_(*t, c*) = *f*_1_(*c*) and *f*_0_(*t, c*) = *f*_0_(*c*), i.e., *AL*(*t, c*) = *AL*(*c*) (ALnoT, AL does not accumulate with age).

### Sensitivity analyses

The LLFS is a family-based study that contains related individuals. Currently available JM tools allowing analyses of related individuals (R-packages *merlin* and *rstanarm*) were not usable for our applications because of technical issues. SPM tools for related samples currently do not exist. Therefore, we used the available tools for unrelated individuals. To test whether this could affect our results, we performed sensitivity analyses implementing the “familial bootstrap” approach [35]. Specifically, we collected estimates of the respective models (JM and SPM) from 100 bootstrap samples constructed from data on the families generated (with replacement) from the original analytic sample (note, that, even though the number of families in each generated sample was the same, the numbers of individuals were different). Then, we computed relevant quantities from all 100 samples (e.g., medians of hazard ratios of the association parameter *α* [Eq. 1] in JM, along with the range of the hazard ratios). The respective estimates are provided in **Supplementary Materials** and discussed in **RESULTS**.

## Data Availability

The Long Life Family Study (LLFS) data used in this study were provided by the LLFS Data Management and Coordinating Center (DMCC), Washington University, St. Louis. The database of Genotypes and Phenotypes (dbGaP) also provides access to phenotypic and genetic LLFS data (dbGaP Study Accession: phs000397.v3.p3). Metabolomic data will become available on the ELITE portal (https://eliteportal.synapse.org/).

## ABBREVIATIONS

AL: allostatic load
*APOE*: apolipoprotein E
CI: confidence interval(s)
H0’s: null hypotheses
HR: hazard ratio
JM: joint model(s)
LLFS: Long Life Family Study
LPC: lysophosphatidylcholine(s)
PCs: principal components
SPM: stochastic process model(s)

## AUTHOR CONTRIBUTIONS

KGA: conceived and designed the study, contributed to statistical analyses, and wrote the manuscript; OB: prepared phenotypic analytic files, performed statistical analyses, prepared tables and figures, and contributed to writing Materials and Methods section; SVU, AK, ES, *Y*G, AI*Y*, MAP: provided feedback on analyses and results; MAP: provided funding; MSH, GJP: generated and cleaned metabolomics data. All authors provided critical comments on the final version of the manuscript.

## ETHICAL STATEMENT AND CONSENT

Written informed consent was obtained from all subjects following protocols approved in the US by the respective field center’s Institutional Review Boards (IRBs) and, in Denmark, by the Regional Committees on Health Research Ethics for Southern Denmark. In this paper, we performed secondary analyses of LLFS data collected at all field centers. This study was approved by the Duke University Health System IRB.

## REFERENCES

1. Therneau T and Grambsch P. (2000). Modeling Survival Data: Extending the Cox Model. (New York: Springer-Verlag).

2. Prentice RL. Covariate measurement errors and parameter estimation in a failure time regression model. Biometrika. 1982; 69(2):331–342.

3. Sweeting MJ and Thompson SG. Joint modelling of longitudinal and time-to-event data with application to predicting abdominal aortic aneurysm growth and rupture. Biom J. 2011; 53(5):750–763.

4. Rizopoulos D. (2012). Joint Models for Longitudinal and Time-to-Event Data With Applications in R. (Boca Raton, FL: Chapman and Hall/CRC).

5. Elashoff RM, Li G and Li N. (2016). Joint Modeling of Longitudinal and Time-to-Event Data. (Boca Raton, FL: CRC Press).

6. Klimczak-Tomaniak D, de Bakker M, Bouwens E, Akkerhuis KM, Baart S, Rizopoulos D, Mouthaan H, van Ramshorst J, Germans T, Constantinescu A, Manintveld O, Umans V, Boersma E, et al. Dynamic personalized risk prediction in chronic heart failure patients: a longitudinal, clinical investigation of 92 biomarkers (Bio-SHiFT study). Scientific Reports. 2022; 12(1):2795.

7. de Bakker M, Petersen TB, Rueten-Budde AJ, Akkerhuis KM, Umans VA, Brugts JJ, Germans T, Reinders MJT, Katsikis PD, van der Spek PJ, Ostroff R, She R, Lanfear D, et al. Machine learning–based biomarker profile derived from 4210 serially measured proteins predicts clinical outcome of patients with heart failure. European Heart Journal - Digital Health. 2023; 4(6):444–454.

8. de Bakker M, Loncq de Jong M, Petersen T, de Lange I, Akkerhuis KM, Umans VA, Rizopoulos D, Boersma E, Brugts JJ and Kardys I. Sex-specific cardiovascular protein levels and their link with clinical outcome in heart failure. ESC Heart Failure. 2024; 11(1):594–600.

9. Woodbury MA and Manton KG. A random-walk model of human mortality and aging. Theoretical Population Biology. 1977; 11(1):37–48.

10. Yashin AI, Manton KG and Vaupel JW. Mortality and aging in a heterogeneous population: A stochastic process model with observed and unobserved variables. Theoretical Population Biology. 1985; 27(2):154–175.

11. Yashin AI, Manton KG and Stallard E. The propagation of uncertainty in human mortality processes operating in stochastic environments. Theoretical Population Biology. 1989; 35(2):119–141.

12. Yashin AI, Arbeev KG, Akushevich I, Kulminski A, Akushevich L and Ukraintseva SV. Stochastic model for analysis of longitudinal data on aging and mortality. Math Biosci. 2007; 208(2):538–551.

13. Arbeev KG, Ukraintseva SV, Akushevich I, Kulminski AM, Arbeeva LS, Akushevich L, Culminskaya IV and Yashin AI. Age trajectories of physiological indices in relation to healthy life course. Mech Ageing Dev. 2011; 132(3):93–102.

14. Arbeev KG, Akushevich I, Kulminski AM, Arbeeva LS, Akushevich L, Ukraintseva SV, Culminskaya IV and Yashin AI. Genetic model for longitudinal studies of aging, health, and longevity and its potential application to incomplete data. Journal of Theoretical Biology. 2009; 258(1):103–111.

15. Yashin AI, Arbeev KG, Akushevich I, Kulminski A, Ukraintseva SV, Stallard E and Land KC. The quadratic hazard model for analyzing longitudinal data on aging, health, and the life span. Physics of Life Reviews. 2012; 9(2):177–188.

16. Arbeev KG, Ukraintseva SV and Yashin AI. Dynamics of biomarkers in relation to aging and mortality. Mech Ageing Dev. 2016; 156:42–54.

17. Arbeev KG, Bagley O, Yashkin AP, Duan H, Akushevich I, Ukraintseva SV and Yashin AI. Understanding Alzheimer’s disease in the context of aging: Findings from applications of stochastic process models to the Health and Retirement Study. Mech Ageing Dev. 2023; 211:111791.

18. Arbeev KG, Ukraintseva SV, Bagley O, Zhbannikov IY, Cohen AA, Kulminski AM and Yashin AI. “Physiological dysregulation” as a promising measure of robustness and resilience in studies of aging and a new indicator of preclinical disease. J Gerontol A Biol Sci Med Sci. 2019; 74(4):462–468.

19. Yashin AI, Arbeev KG, Akushevich I, Ukraintseva SV, Kulminski A, Arbeeva LS and Culminskaya I. Exceptional survivors have lower age trajectories of blood glucose: lessons from longitudinal data. Biogerontology. 2010; 11(3):257–265.

20. Yashin AI, Arbeev KG, Kulminski A, Akushevich I, Akushevich L and Ukraintseva SV. Health decline, aging and mortality: how are they related? Biogerontology. 2007; 8(3):291–302.

21. Yashin AI, Arbeev KG, Ukraintseva SV, Akushevich I and Kulminski A. Patterns of aging related changes on the way to 100: An approach to studying aging, mortality, and longevity from longitudinal data. N Amer Actuarial J. 2012; 16(4):403–433.

22. Yashin AI, Arbeev KG, Wu D, Arbeeva L, Kulminski A, Kulminskaya I, Akushevich I and Ukraintseva SV. How genes modulate patterns of aging-related changes on the way to 100: Biodemographic models and methods in genetic analyses of longitudinal data. N Amer Actuarial J. 2016; 20(3):201–232.

23. Yashin AI, Ukraintseva SV, Arbeev KG, Akushevich I, Arbeeva LS and Kulminski AM. Maintaining physiological state for exceptional survival: What is the normal level of blood glucose and does it change with age? Mech Ageing Dev. 2009; 130(9):611–618.

24. Wojczynski MK, Jiuan Lin S, Sebastiani P, Perls TT, Lee J, Kulminski A, Newman A, Zmuda JM, Christensen K, Province MA and on behalf of the Long Life Family Study. NIA Long Life Family Study: Objectives, design, and heritability of cross-sectional and longitudinal phenotypes. The Journals of Gerontology: Series A. 2022; 77(4):717–727.

25. Law SH, Chan ML, Marathe GK, Parveen F, Chen CH and Ke LY. An Updated Review of Lysophosphatidylcholine Metabolism in Human Diseases. International Journal of Molecular Sciences. 2019; 20(5):1149.

26. Barupal DK, Baillie R, Fan S, Saykin AJ, Meikle PJ, Arnold M, Nho K, Fiehn O, Kaddurah-Daouk R, Alzheimer’s Disease Neuroimaging I and Alzheimer Disease Metabolomics C. Sets of coregulated serum lipids are associated with Alzheimer’s disease pathophysiology. Alzheimer’s & Dementia: Diagnosis, Assessment & Disease Monitoring. 2019; 11(1):619–627.

27. Diray-Arce J, Conti MG, Petrova B, Kanarek N, Angelidou A and Levy O. (2020). Integrative Metabolomics to Identify Molecular Signatures of Responses to Vaccines and Infections. Metabolites.

28. Semba RD. Perspective: The Potential Role of Circulating Lysophosphatidylcholine in Neuroprotection against Alzheimer Disease. Advances in Nutrition. 2020; 11(4):760–772.

29. Peña-Bautista C, Álvarez-Sánchez L, Roca M, García-Vallés L, Baquero M and Cháfer-Pericás C. (2022). Plasma Lipidomics Approach in Early and Specific Alzheimer’s Disease Diagnosis. Journal of Clinical Medicine.

30. Dorninger F, Moser AB, Kou J, Wiesinger C, Forss-Petter S, Gleiss A, Hinterberger M, Jungwirth S, Fischer P and Berger J. Alterations in the Plasma Levels of Specific Choline Phospholipids in Alzheimer’s Disease Mimic Accelerated Aging. J Alzheimer’s Dis. 2018; 62:841–854.

31. Liu D, Aziz NA, Landstra EN and Breteler MMB. The lipidomic correlates of epigenetic aging across the adult lifespan: A population-based study. Aging Cell. 2023; 22(9):e13934.

32. Knuplez E and Marsche G. An Updated Review of Pro-and Anti-Inflammatory Properties of Plasma Lysophosphatidylcholines in the Vascular System. International Journal of Molecular Sciences. 2020; 21(12):4501.

33. Rizopoulos D. JM: An R Package for the Joint Modelling of Longitudinal and Time-to-Event Data. Journal of Statistical Software. 2010; 35(9):1–33.

34. Arbeev KG, Bagley O, Ukraintseva SV, Wu D, Duan H, Kulminski AM, Stallard E, Christensen K, Lee JH, Thyagarajan B, Zmuda JM and Yashin AI. Genetics of physiological dysregulation: findings from the long life family study using joint models. Aging. 2020; 12(7):5920–5947.

35. Borecki IB and Province MA. Genetic and Genomic Discovery Using Family Studies. Circulation. 2008; 118(10):1057–1063.

36. Wulfsohn MS and Tsiatis AA. A joint model for survival and longitudinal data measured with error. Biometrics. 1997; 53(1):330–339.

37. Henderson R, Diggle P and Dobson A. Joint modelling of longitudinal measurements and event time data. Biostatistics. 2000; 1(4):465–480.

38. Henderson R, Diggle P and Dobson A. Identification and efficacy of longitudinal markers for survival. Biostatistics. 2002; 3(1):33–50.

39. Johnson JD and Alejandro EU. Control of pancreatic β-cell fate by insulin signaling: The sweet spot hypothesis. Cell Cycle. 2008; 7(10):1343–1347.

40. Lee JH, Han K and Huh JH. The sweet spot: fasting glucose, cardiovascular disease, and mortality in older adults with diabetes: a nationwide population-based study. Cardiovascular Diabetology. 2020; 19(1):44.

41. Vishnyakova O, Song X, Rockwood K, Elliott LT and Brooks-Wilson A. Physiological phenotypes have optimal values relevant to healthy aging: sweet spots deduced from the Canadian Longitudinal Study on Aging. Geroscience. 2024; 46(2):1589–1605.

42. Ukraintseva S, Arbeev K, Duan M, Akushevich I, Kulminski A, Stallard E and Yashin A. Decline in biological resilience as key manifestation of aging: Potential mechanisms and role in health and longevity. Mech Ageing Dev. 2021; 194:111418.

43. Li X, Yin Z, Yan W, Wang M, Chang C, Guo C, Xue L, Zhou Q and Sun Y. Association between Changes in Plasma Metabolism and Clinical Outcomes of Sepsis. Emergency Medicine International. 2023; 2023:2590115.

44. Trovato FM, Zia R, Napoli S, Wolfer K, Huang X, Morgan PE, Husbyn H, Elgosbi M, Lucangeli M, Miquel R, Wilson I, Heaton ND, Heneghan MA, et al. Dysregulation of the Lysophosphatidylcholine/Autotaxin/Lysophosphatidic Acid Axis in Acute-on-Chronic Liver Failure Is Associated With Mortality and Systemic Inflammation by Lysophosphatidic Acid–Dependent Monocyte Activation. Hepatology. 2021; 74(2):907–925.

45. Banoei MM, Vogel HJ, Weljie AM, Yende S, Angus DC and Winston BW. Plasma lipid profiling for the prognosis of 90-day mortality, in-hospital mortality, ICU admission, and severity in bacterial community-acquired pneumonia (CAP). Crit Care. 2020; 24(1):461.

46. Trovato FM, Zia R, Artru F, Mujib S, Jerome E, Cavazza A, Coen M, Wilson I, Holmes E, Morgan P, Singanayagam A, Bernsmeier C, Napoli S, et al. Lysophosphatidylcholines modulate immunoregulatory checkpoints in peripheral monocytes and are associated with mortality in people with acute liver failure. J Hepatol. 2023; 78(3):558–573.

47. Tian Q, Adam MG, Ozcariz E, Fantoni G, Shehadeh NM, Turek LM, Collingham VL, Kaileh M, Moaddel R and Ferrucci L. Human Metabolome Reference Database in a Biracial Cohort across the Adult Lifespan. Metabolites. 2023; 13(5):591.

48. Tian Q, Mitchell BA, Zampino M and Ferrucci L. Longitudinal associations between blood lysophosphatidylcholines and skeletal muscle mitochondrial function. GeroScience. 2022; 44(4):2213–2221.

49. Semba RD, Zhang P, Adelnia F, Sun K, Gonzalez-Freire M, Salem Jr N, Brennan N, Spencer RG, Fishbein K, Khadeer M, Shardell M, Moaddel R and Ferrucci L. Low plasma lysophosphatidylcholines are associated with impaired mitochondrial oxidative capacity in adults in the Baltimore Longitudinal Study of Aging. Aging Cell. 2019; 18(2):e12915.

50. Tian Q, Shardell MD, Kuo P-L, Tanaka T, Simonsick EM, Moaddel R, Resnick SM and Ferrucci L. Plasma metabolomic signatures of dual decline in memory and gait in older adults. GeroScience. 2023; 45(4):2659–2667.

51. Hornburg D, Wu S, Moqri M, Zhou X, Contrepois K, Bararpour N, Traber GM, Su B, Metwally AA, Avina M, Zhou W, Ubellacker JM, Mishra T, et al. Dynamic lipidome alterations associated with human health, disease and ageing. Nature Metabolism. 2023; 5(9):1578–1594.

52. Mohammadzadeh Honarvar N, Zarezadeh M, Molsberry SA and Ascherio A. Changes in plasma phospholipids and sphingomyelins with aging in men and women: A comprehensive systematic review of longitudinal cohort studies. Ageing Research Reviews. 2021; 68:101340.

53. He L, Zhbannikov I, Arbeev KG, Yashin AI and Kulminski AM. A genetic stochastic process model for genome-wide joint analysis of biomarker dynamics and disease susceptibility with longitudinal data. Genetic Epidemiology. 2017; 41(7):620–635.

54. Hickey GL, Philipson P, Jorgensen A and Kolamunnage-Dona R. Joint modelling of time-to-event and multivariate longitudinal outcomes: recent developments and issues. BMC Medical Research Methodology. 2016; 16:15.

55. Arbeev KG, Cohen AA, Arbeeva LS, Milot E, Stallard E, Kulminski AM, Akushevich I, Ukraintseva S, Christensen K and Yashin AI. Optimal versus realized trajectories of physiological dysregulation in aging and their relation to sex-specific mortality risk. Frontiers in Public Health. 2016; 4:article 3.

56. Sebastiani P, Hadley EC, Province M, Christensen K, Rossi W, Perls TT and Ash AS. A Family Longevity Selection Score: Ranking sibships by their longevity, size, and availability for study. American Journal of Epidemiology. 2009; 170(12):1555–1562.

57. Elo IT, Mykyta L, Sebastiani P, Christensen K, Glynn NW and Perls T. Age Validation in the Long Life Family Study Through a Linkage to Early-Life Census Records. J Gerontol B Psychol Sci Soc Sci. 2013; 68(4):580–585.

58. Stancliffe E, Schwaiger-Haber M, Sindelar M, Murphy MJ, Soerensen M and Patti GJ. An Untargeted Metabolomics Workflow that Scales to Thousands of Samples for Population-Based Studies. Analytical Chemistry. 2022; 94(50):17370–17378.

59. Mahieu NG, Spalding JL, Gelman SJ and Patti GJ. Defining and Detecting Complex Peak Relationships in Mass Spectral Data: The Mz.unity Algorithm. Anal Chem. 2016; 88(18):9037–9046.

60. Cho K, Schwaiger-Haber M, Naser FJ, Stancliffe E, Sindelar M and Patti GJ. Targeting unique biological signals on the fly to improve MS/MS coverage and identification efficiency in metabolomics. Anal Chim Acta. 2021; 1149:338210.

61. Stancliffe E, Schwaiger-Haber M, Sindelar M and Patti GJ. DecoID improves identification rates in metabolomics through database-assisted MS/MS deconvolution. Nat Methods. 2021; 18(7):779–787.

62. Jin Z, Kang J and Yu T. Missing value imputation for LC-MS metabolomics data by incorporating metabolic network and adduct ion relations. Bioinformatics. 2018; 34(9):1555–1561.

63. Fan S, Kind T, Cajka T, Hazen SL, Tang WHW, Kaddurah-Daouk R, Irvin MR, Arnett DK, Barupal DK and Fiehn O. Systematic Error Removal Using Random Forest for Normalizing Large-Scale Untargeted Lipidomics Data. Analytical Chemistry. 2019; 91(5):3590–3596.

64. Fahy E and Subramaniam S. RefMet: a reference nomenclature for metabolomics. Nat Methods. 2020; 17(12):1173–1174.

65. Rizopoulos D. Fast fitting of joint models for longitudinal and event time data using a pseudo-adaptive Gaussian quadrature rule. Comput Stat Data Anal. 2012; 56(3):491–501.

66. McEwen BS and Wingfield JC. The concept of allostasis in biology and biomedicine. Hormones and Behavior. 2003; 43(1):2–15.

67. Ukraintseva S, Yashin AI and Arbeev KG. Resilience versus robustness in aging. J Gerontol A Biol Sci Med Sci. 2016; 71(11):1533–1534.

68. Strehler B. (1962). Time, Cells, and Aging. (London: Academic Press).

69. Strehler BL and Mildvan AS. General theory of mortality and aging. Science. 1960; 132(3418):14–21.

70. López-Otín C, Blasco MA, Partridge L, Serrano M and Kroemer G. Hallmarks of aging: An expanding universe. Cell. 2023; 186(2):243–278.

71. Sterling P and Eyer J. (1988). Allostasis: A New Paradigm to Explain Arousal Pathology. In: Fisher S and Reason J, eds. Handbook of Life Stress, Cognition and Health. (New York: John Wiley & Sons), pp. 629–649.

72. McEwen BS and Stellar E. Stress and the individual. Mechanisms leading to disease. Arch Intern Med. 1993; 153(18):2093–2101.

73. McEwen BS. (1998). Stress, adaptation, and disease. Allostasis and allostatic load. In: McCann SM, Lipton JM, Sternberg EM, Chrousos GP, Gold PW and Smith CC, eds. Neuroimmunomodulation: Molecular Aspects, Integrative Systems, and Clinical Advances. (New York: New York Acad Sciences), pp. 33–44.

