## Supplementary Material for "Methods for joint modelling of longitudinal omics data and time-to-event outcomes: Applications to lysophosphatidylcholines in connection to aging and mortality in the Long Life Family Study"

**Supplementary Figures**

| 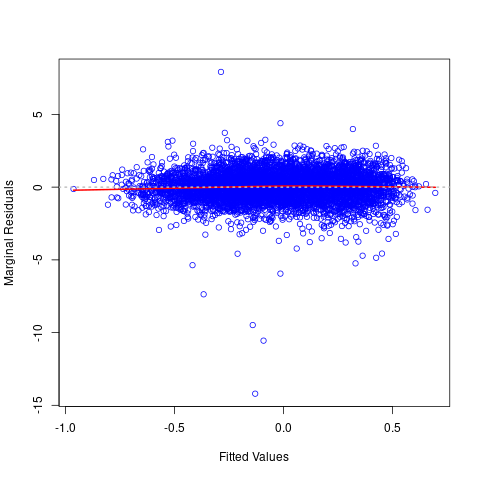  **a** | 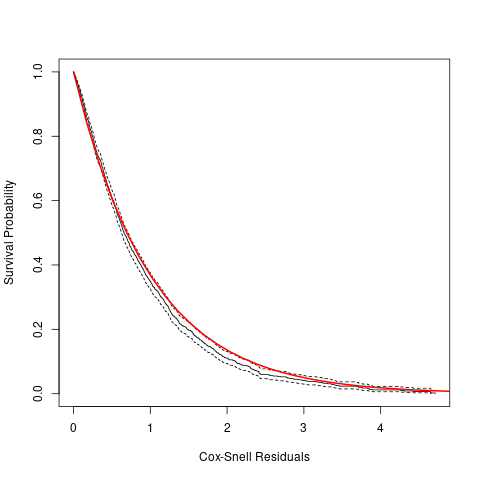  **b** |
| --- | --- |

**Figure S1:** **Diagnostic plots assessing the goodness-of-fit and assumptions of joint models in applications to LPC 15:0/0:0.** a) Plot of the standardized marginal residuals versus the corresponding fitted values for the longitudinal outcome (LPC 15:0/0:0). The red solid line denotes the fit of the loess smoother. b) Residuals analysis for the survival outcome by assessing the overall fit of the survival submodel using the Cox-Snell residuals. The black solid line denotes the Kaplan-Meier estimate of the survival function of the residuals (with the dashed lines corresponding to the 95% pointwise confidence intervals). The red solid line is the survival function of the unit exponential distribution (this is the distribution in case if the survival submodel is correct).

| 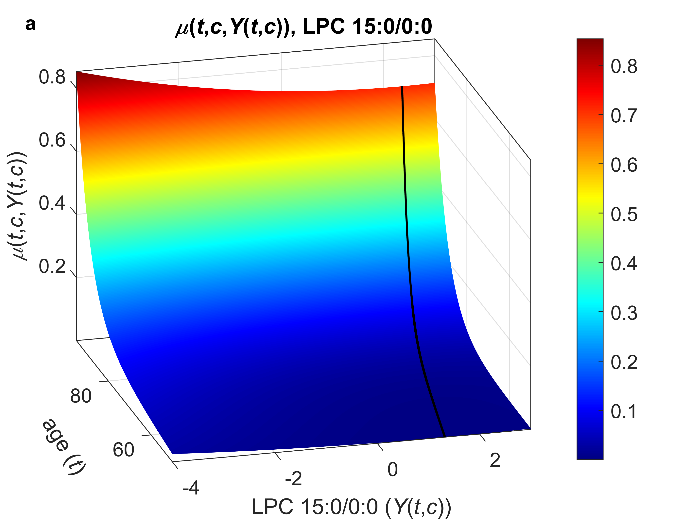 | 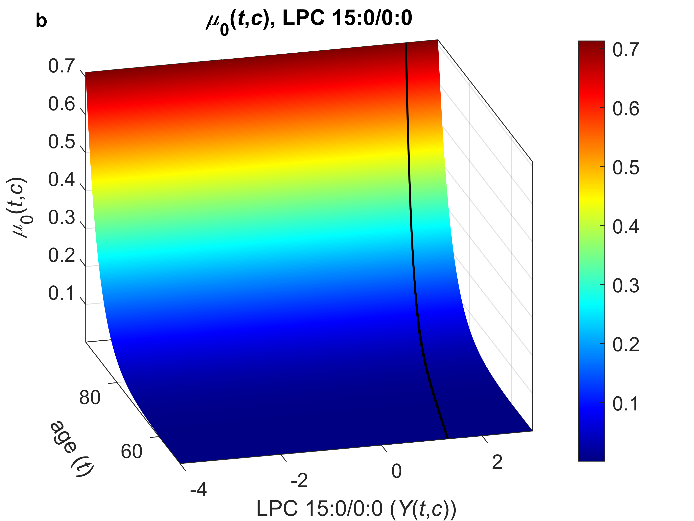 |
| --- | --- |
| 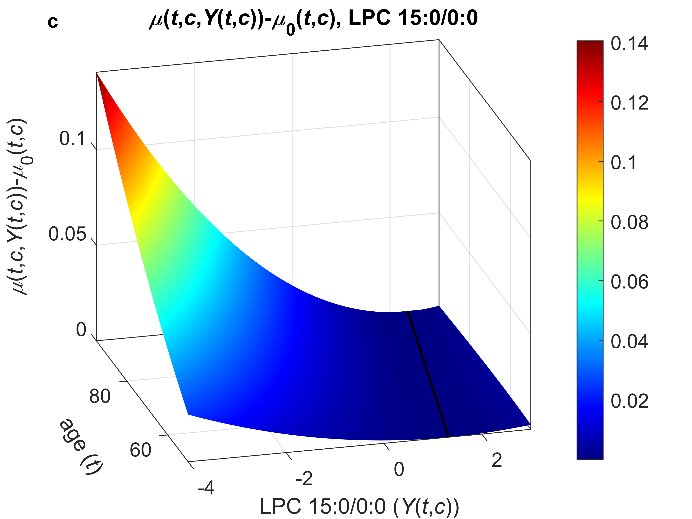 | 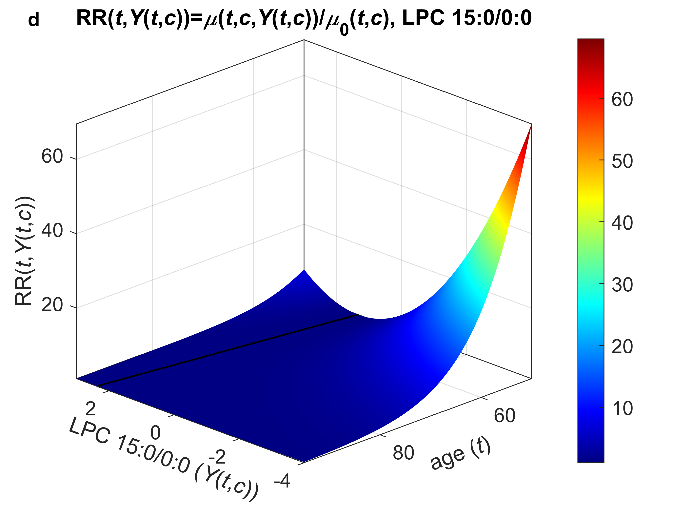 |

**Figure S2: 3D plots illustrating results of applications of stochastic process models to measurements of LPC 15:0/0:0 and mortality data in the LLFS.** a) Estimates of the total mortality rate as a function of age (*t*) and the metabolite ($Y\left( t, c \right)$); b) Estimates of the baseline mortality rate $\mu_{0}\left( t, c \right)$ (which does not depend on $Y\left( t, c \right)$); c) Estimates of the quadratic term in mortality rate ($\mu\left( t,c,Y\left( t,c \right) \right)- \mu_{0}\left( t, c \right)$) for different ages (*t*) and values of the metabolite ($Y\left( t, c \right)$); d) Estimates of the relative risk $RR\left( t,Y\left( t,c \right) \right)={\mu\left( t,c,Y\left( t,c \right) \right)}/{\mu_{0}\left( t, c \right)}$. The estimates are shown for the values of the covariate “SexM” corresponding to females (i.e., SexM=0, see **Notes** under **Table S6**). Black line denotes the optimal trajectory $f_{0}\left( t,c \right)$. LPC values were log-transformed and standardized (see **Data**).

**Supplementary Tables**

**Table S1:** Characteristics of the Long Life Family Study metabolomics sample (batch 6, released on October 25, 2023)

| **Characteristics** | **Field Center** | | | | **Total Sample** |
| --- | --- | --- | --- | --- | --- |
|  | **BU** | **NY** | **PT** | **DK** |  |
| Number of families | 242 | 262 | 221 | 78 | 583 |
| Number of participants at any visit | 1,280 | 870 | 1,192 | 1,239 | 4,581 |
| Number of participants at visit 1 | 1,173 | 713 | 1,139 | 1,196 | 4,221 |
| Number of participants at visit 2 | 683 | 487 | 587 | 798 | 2,555 |
| Number of participants with genetic PCs | 1,268 | 842 | 1,181 | 1,229 | 4,520 |
| Number (%) of deceased participants | 409  (32.0%) | 327  (37.6%) | 445  (37.3%) | 370  (29.9%) | 1,551  (33.9%) |
| Follow-up period (years) (mean ± SD [range]) | 10.0 ± 4.5  [0.00, 17.00] | 9.6 ± 4.2  [0.00, 17.00] | 10.4 ± 4.3  [0.29, 17.00] | 11.1 ± 5.1  [0.00, 17.00] | 10.3 ± 4.6  [0.00, 17.00] |
| Age at baseline (mean ± SD [range]) | 69.6 ± 16.0  [32, 110] | 73.6 ± 16.1  [24, 108] | 71.1 ± 15.9  [36, 104] | 67.3 ± 14.3  [36, 104] | 70.0 ± 15.7  [24, 110] |
| Whites (%) | 99.2% | 98.2% | 99.7% | 99.0% | 99.1% |
| Females (%) | 55.6% | 54.6% | 55.9% | 54.3% | 55.1% |
| Low educated participants (below high school) (%) | 5.9% | 7.5% | 7.1% | 27.7% | 12.4% |
| Smokers (smoked >100 cigarettes in lifetime) (%) | 41.4% | 45.6% | 35.7% | 49.2% | 42.8% |
| *APOE* ɛ4 allele carriers (%) | 13.8% | 17.4% | 17.0% | 25.3% | 18.4% |
| Medication use: angina (%) | 32.0% | 30.5% | 32.9% | 23.5% | 29.6% |
| Medication use: anti-diabetic (%) | 6.9% | 7.5% | 9.1% | 5.5% | 7.2% |
| Medication use: anti-hypertensive (%) | 50.4% | 52.3% | 55.2% | 42.0% | 49.7% |
| Medication use: lipid-lowering (%) | 35.0% | 43.3% | 39.1% | 21.1% | 33.9% |

**Notes:** a) Genetic PCs were computed from LLFS whole-genome sequencing data; b) Number of missing data: race – 20, education – 10, smoking – 18, *APOE* – 257, angina medications – 273, anti-diabetic drugs – 273, anti-hypertensive drugs – 273, lipid-lowering drugs – 273, other variables listed in the table have no missing values; c) The numbers shown in “Number of deceased participants” and “Follow-up period” correspond to the LLFS data release used in this paper (see **Data**). Abbreviations: BU – Boston, DK – Denmark, NY – New York, PT – Pittsburgh, SD – standard deviation.

**Table S2:** Results of applications of joint models to LPC 15:0/0:0 and mortality data in the LLFS: Estimates of parameters in the longitudinal and survival sub-models

| **Sex** | **Variable** | **Longitudinal** | | **Survival** | | |
| --- | --- | --- | --- | --- | --- | --- |
|  |  | **Beta** | **95% CI** | **Beta** | **HR** | **95% CI for HR** |
| Total | LPC |  |  | -0.335 | 0.715 | (0.649, 0.788) |
|  | Intercept_longit | 1.034 | (0.883, 1.185) |  |  |  |
|  | AgeV1 | -0.012 | (-0.014, -0.010) | 0.123 | 1.131 | (1.124, 1.137) |
|  | TimeV1 | 0.011 | (0.006, 0.017) |  |  |  |
|  | SexM | -0.086 | (-0.143, -0.029) | 0.253 | 1.288 | (1.154, 1.437) |
|  | IsDK | -0.304 | (-0.372, -0.235) | 0.040 | 1.041 | (0.900, 1.204) |
|  | LowEduc | -0.063 | (-0.158, 0.031) | 0.093 | 1.098 | (0.954, 1.262) |
|  | Smoke100 | -0.083 | (-0.139, -0.026) | 0.145 | 1.156 | (1.035, 1.291) |
|  | APOE4 | -0.019 | (-0.090, 0.051) | 0.193 | 1.213 | (1.049, 1.401) |
|  | MedsDiab | -0.165 | (-0.272, -0.059) |  |  |  |
|  | MedsHtn | -0.016 | (-0.077, 0.045) |  |  |  |
|  | MedsLipid | -0.218 | (-0.275, -0.161) |  |  |  |
|  | MedsNitro | -0.054 | (-0.120, 0.012) |  |  |  |
|  | PC1 |  |  | 0.108 | 1.114 | (0.937, 1.325) |
|  | PC2 |  |  | 0.057 | 1.059 | (0.897, 1.251) |
| Females | LPC |  |  | -0.300 | 0.741 | (0.650, 0.844) |
|  | Intercept_longit | 0.907 | (0.705, 1.109) |  |  |  |
|  | AgeV1 | -0.010 | (-0.013, -0.008) | 0.123 | 1.131 | (1.122, 1.139) |
|  | TimeV1 | 0.013 | (0.007, 0.019) |  |  |  |
|  | IsDK | -0.303 | (-0.397, -0.209) | -0.006 | 0.994 | (0.808, 1.223) |
|  | LowEduc | -0.005 | (-0.127, 0.117) | 0.103 | 1.108 | (0.916, 1.341) |
|  | Smoke100 | -0.067 | (-0.145, 0.011) | 0.132 | 1.141 | (0.971, 1.340) |
|  | APOE4 | -0.073 | (-0.168, 0.022) | 0.201 | 1.223 | (0.997, 1.501) |
|  | MedsDiab | -0.215 | (-0.364, -0.066) |  |  |  |
|  | MedsHtn | -0.033 | (-0.113, 0.046) |  |  |  |
|  | MedsLipid | -0.193 | (-0.269, -0.116) |  |  |  |
|  | MedsNitro | -0.022 | (-0.113, 0.068) |  |  |  |
|  | PC1 |  |  | 0.169 | 1.184 | (0.928, 1.511) |
|  | PC2 |  |  | 0.153 | 1.165 | (0.922, 1.472) |
| Males | LPC |  |  | -0.267 | 0.766 | (0.667, 0.879) |
|  | Intercept_longit | 1.115 | (0.888, 1.343) |  |  |  |
|  | AgeV1 | -0.014 | (-0.017, -0.011) | 0.123 | 1.131 | (1.122, 1.141) |
|  | TimeV1 | 0.010 | (0.001, 0.019) |  |  |  |
|  | IsDK | -0.315 | (-0.415, -0.215) | 0.097 | 1.102 | (0.898, 1.353) |
|  | LowEduc | -0.181 | (-0.333, -0.029) | 0.091 | 1.095 | (0.888, 1.351) |
|  | Smoke100 | -0.081 | (-0.164, 0.002) | 0.168 | 1.183 | (1.017, 1.376) |
|  | APOE4 | 0.046 | (-0.058, 0.150) | 0.191 | 1.211 | (0.988, 1.485) |
|  | MedsDiab | -0.098 | (-0.247, 0.051) |  |  |  |
|  | MedsHtn | 0.011 | (-0.085, 0.106) |  |  |  |
|  | MedsLipid | -0.247 | (-0.332, -0.162) |  |  |  |
|  | MedsNitro | -0.100 | (-0.198, -0.002) |  |  |  |
|  | PC1 |  |  | 0.052 | 1.053 | (0.865, 1.280) |
|  | PC2 |  |  | -0.037 | 0.964 | (0.795, 1.169) |

**Notes:** **Variable** – LPC: true (unobserved) value of the metabolite (in **Survival** only), Intercept_longit: intercept (in **Longitudinal** only), AgeV1: age at visit 1, TimeV1: time since visit 1 (in **Longitudinal** only), SexM: sex (1 – male, 0 – female), IsDK: country (1 – Denmark, 0 – USA), LowEduc: low education (1 – below high school, 0 – otherwise), Smoke100: smoking (1 – smoked 100 cigarettes in lifetime, 0 - otherwise), APOE4: *APOE* ɛ4 carrier status (1 – carrier, 0 – non-carrier), MedsDiab: diabetes mellitus medications (1 – taking; 0 – not taking) (in **Longitudinal** only), MedsHtn: hypertension medications (1 – taking; 0 – not taking) (in **Longitudinal** only), MedsLipid: lipid lowering medications (1 – taking; 0 – not taking) (in **Longitudinal** only), MedsNitro: angina medications (1 – taking; 0 – not taking) (in **Longitudinal** only), PC1, PC2: principal components computed from LLFS whole-genome sequencing data (in **Survival** only); **Longitudinal** – estimates for the longitudinal sub-model of JM (modeling the metabolite); **Survival** – estimates for the survival sub-model of JM (modeling mortality rate); **Beta** – estimates of regression parameters for **Variable** in respective sub-models; **HR** – hazard ratios computed from **Beta** in **Survival** (HRs are for a unit increase in all variables except PCs where they are for an increase by a standard deviation); **95% CI –** respective 95% confidence intervals. The joint models were estimated using R-package *JM*. LPC values were log-transformed and standardized (see **Data**).

**Table S3:** Results of applications of Cox models to measurements of LPC species and mortality data in the LLFS: Estimates of the association parameter for the metabolite

| **Metabolite** | **Total** | | **Females** | | **Males** | |
| --- | --- | --- | --- | --- | --- | --- |
|  | **Alpha** | **HR (95% CI)** | **Alpha** | **HR (95% CI)** | **Alpha** | **HR (95% CI)** |
| LPC 0:0/16:0 | -0.091 | **0.913 (0.868,0.960)** | -0.123 | **0.884 (0.816,0.957)** | -0.075 | **0.928 (0.867,0.994)** |
| LPC 0:0/16:1 | 0.007 | 1.007 (0.952,1.065) | -0.017 | 0.983 (0.907,1.065) | 0.025 | 1.025 (0.947,1.110) |
| LPC 0:0/18:0 | -0.024 | 0.976 (0.924,1.031) | -0.04 | 0.961 (0.890,1.038) | -0.012 | 0.988 (0.912,1.070) |
| LPC 0:0/18:1 | -0.06 | **0.942 (0.889,0.998)** | -0.062 | 0.94 (0.864,1.023) | -0.064 | 0.938 (0.866,1.015) |
| LPC 0:0/18:2 | -0.097 | **0.908 (0.857,0.963)** | -0.071 | 0.931 (0.856,1.013) | -0.128 | **0.88 (0.812,0.954)** |
| LPC 0:0/20:3 | -0.028 | 0.972 (0.922,1.025) | -0.045 | 0.956 (0.887,1.030) | -0.02 | 0.98 (0.909,1.057) |
| LPC 0:0/20:4 | -0.078 | **0.925 (0.874,0.978)** | -0.103 | **0.902 (0.834,0.976)** | -0.06 | 0.942 (0.868,1.022) |
| LPC 0:0/22:6 | -0.119 | **0.888 (0.839,0.940)** | -0.176 | **0.839 (0.771,0.913)** | -0.077 | 0.926 (0.856,1.002) |
| LPC 14:0/0:0 | -0.081 | **0.922 (0.875,0.971)** | -0.113 | **0.893 (0.824,0.967)** | -0.063 | 0.939 (0.874,1.009) |
| LPC 15:0/0:0 | -0.211 | **0.81 (0.769,0.853)** | -0.218 | **0.804 (0.742,0.871)** | -0.211 | **0.81 (0.756,0.868)** |
| LPC 16:0/0:0 | -0.089 | **0.915 (0.874,0.957)** | -0.143 | **0.867 (0.800,0.940)** | -0.067 | **0.935 (0.881,0.993)** |
| LPC 16:1/0:0 | -0.012 | 0.988 (0.934,1.046) | -0.028 | 0.972 (0.898,1.052) | -0.003 | 0.997 (0.919,1.082) |
| LPC 17:0/0:0 | -0.109 | **0.897 (0.852,0.944)** | -0.127 | **0.881 (0.818,0.949)** | -0.097 | **0.908 (0.844,0.976)** |
| LPC 18:0/0:0 | 0.005 | 1.005 (0.949,1.064) | -0.013 | 0.987 (0.913,1.067) | 0.023 | 1.023 (0.939,1.114) |
| LPC 18:1/0:0 | -0.084 | **0.919 (0.877,0.964)** | -0.119 | **0.888 (0.819,0.964)** | -0.071 | **0.931 (0.876,0.990)** |
| LPC 18:2/0:0 | -0.082 | **0.921 (0.880,0.963)** | -0.102 | **0.903 (0.827,0.987)** | -0.079 | **0.924 (0.876,0.974)** |
| LPC 18:3/0:0 | -0.078 | **0.925 (0.873,0.979)** | -0.065 | 0.937 (0.864,1.016) | -0.108 | **0.898 (0.827,0.975)** |
| LPC 20:2/0:0 | -0.083 | **0.92 (0.873,0.970)** | -0.083 | **0.92 (0.853,0.993)** | -0.092 | **0.912 (0.848,0.980)** |
| LPC 20:3/0:0 | -0.095 | **0.909 (0.862,0.959)** | -0.114 | **0.892 (0.825,0.964)** | -0.087 | **0.917 (0.852,0.988)** |
| LPC 20:4/0:0 | -0.092 | **0.912 (0.862,0.965)** | -0.108 | **0.898 (0.828,0.975)** | -0.084 | **0.919 (0.849,0.994)** |
| LPC 20:5/0:0 | -0.159 | **0.853 (0.805,0.903)** | -0.218 | **0.804 (0.736,0.877)** | -0.12 | **0.887 (0.820,0.960)** |
| LPC 22:5/0:0 | -0.1 | **0.905 (0.858,0.954)** | -0.14 | **0.869 (0.804,0.939)** | -0.073 | 0.93 (0.864,1.001) |
| LPC 22:6/0:0 | -0.152 | **0.859 (0.812,0.909)** | -0.176 | **0.839 (0.775,0.908)** | -0.136 | **0.873 (0.805,0.947)** |

**Notes:** **Alpha** – estimates of the association parameter for the longitudinal variable (metabolite) in the Cox model; **HR** – hazard ratios (for a unit increase in the log-transformed and standardized metabolite values) computed from the association parameters; **95% CI** – 95% confidence intervals for HRs (highlighted in **bold** are cases where confidence intervals do not contain one). The analyses were performed using R-package *survival*.

**Table S4:** Estimates of parameters in applications of the Cox model to LPC 15:0/0:0 and mortality data in the LLFS

| **Sex** | **Variable** | **Beta** | **HR** | **95% CI for HR** |
| --- | --- | --- | --- | --- |
| Total | LPC | -0.211 | 0.810 | (0.769,0.853) |
|  | AgeV1 | 0.126 | 1.134 | (1.128,1.141) |
|  | SexM | 0.258 | 1.294 | (1.162,1.442) |
|  | IsDK | 0.052 | 1.053 | (0.913,1.215) |
|  | LowEduc | 0.102 | 1.107 | (0.965,1.269) |
|  | Smoke100 | 0.160 | 1.174 | (1.054,1.308) |
|  | APOE4 | 0.183 | 1.201 | (1.041,1.385) |
|  | PC1 | 0.141 | 1.151 | (0.965,1.372) |
|  | PC2 | 0.070 | 1.072 | (0.904,1.271) |
| Females | LPC | -0.218 | 0.804 | (0.742,0.871) |
|  | AgeV1 | 0.126 | 1.134 | (1.126,1.143) |
|  | IsDK | 0.003 | 1.003 | (0.817,1.232) |
|  | LowEduc | 0.110 | 1.116 | (0.925,1.347) |
|  | Smoke100 | 0.126 | 1.134 | (0.967,1.331) |
|  | APOE4 | 0.204 | 1.226 | (1.001,1.501) |
|  | PC1 | 0.203 | 1.225 | (0.961,1.561) |
|  | PC2 | 0.183 | 1.201 | (0.951,1.516) |
| Males | LPC | -0.211 | 0.810 | (0.756,0.868) |
|  | AgeV1 | 0.126 | 1.134 | (1.125,1.144) |
|  | IsDK | 0.099 | 1.104 | (0.903,1.351) |
|  | LowEduc | 0.108 | 1.114 | (0.908,1.367) |
|  | Smoke100 | 0.193 | 1.213 | (1.044,1.408) |
|  | APOE4 | 0.168 | 1.183 | (0.966,1.449) |
|  | PC1 | 0.073 | 1.076 | (0.875,1.325) |
|  | PC2 | -0.043 | 0.958 | (0.779,1.180) |

**Notes:** **Variable** – LPC: observed value of the metabolite, AgeV1: age at visit 1, SexM: sex (1 – male, 0 – female), IsDK: country (1 – Denmark, 0 – USA), LowEduc: low education (1 – below high school, 0 – otherwise), Smoke100: smoking (1 – smoked 100 cigarettes in lifetime, 0 - otherwise), APOE4: *APOE* ɛ4 carrier status (1 – carrier, 0 – non-carrier), PC1, PC2: principal components computed from LLFS whole-genome sequencing data; **Beta** – estimates of regression parameters for **Variable**; **HR** – hazard ratios computed from **Beta** (HRs are for a unit increase in all variables except PCs where they are for an increase by a standard deviation); **95% CI –** respective 95% confidence intervals. The Cox models were estimated using R-package *survival*. LPC values were log-transformed and standardized (see **Data**).

**Table S5:** Results of applications of joint models with random intercept and slope of LPC 15:0/0:0 in mortality rate in sex-specific analyses of the LLFS

| **Model** | **Sex** | **Variable** | **Longitudinal** | | **Survival** | | |
| --- | --- | --- | --- | --- | --- | --- | --- |
|  |  |  | **Beta** | **95% CI** | **Beta** | **HR** | **95% CI for HR** |
| int | Females | LPC_randomint |  |  | -0.367 | 0.693 | (0.586, 0.788) |
|  |  | Intercept_longit | 0.929 | (0.722, 1.144) |  |  |  |
|  |  | AgeV1 | -0.011 | (-0.014, -0.008) | 0.129 | 1.138 | (1.129, 1.148) |
|  |  | TimeV1 | 0.013 | (0.006, 0.020) |  |  |  |
|  |  | IsDK | -0.311 | (-0.402, -0.214) | 0.086 | 1.089 | (0.881, 1.321) |
|  |  | LowEduc | -0.008 | (-0.120, 0.107) | 0.114 | 1.121 | (0.921, 1.372) |
|  |  | Smoke100 | -0.068 | (-0.152, 0.009) | 0.149 | 1.160 | (1.002, 1.394) |
|  |  | APOE4 | -0.072 | (-0.174, 0.033) | 0.233 | 1.262 | (1.032, 1.594) |
|  |  | MedsDiab | -0.188 | (-0.344, -0.058) |  |  |  |
|  |  | MedsHtn | -0.037 | (-0.117, 0.033) |  |  |  |
|  |  | MedsLipid | -0.215 | (-0.293, -0.135) |  |  |  |
|  |  | MedsNitro | 0.017 | (-0.064, 0.112) |  |  |  |
|  |  | PC1 |  |  | 0.186 | 1.204 | (0.996, 1.484) |
|  |  | PC2 |  |  | 0.170 | 1.185 | (0.975, 1.424) |
|  | Males | LPC_randomint |  |  | -0.543 | 0.581 | (0.458, 0.704) |
|  |  | Intercept_longit | 1.116 | (0.878, 1.342) |  |  |  |
|  |  | AgeV1 | -0.014 | (-0.017, -0.011) | 0.132 | 1.141 | (1.132, 1.153) |
|  |  | TimeV1 | 0.011 | (0.002, 0.021) |  |  |  |
|  |  | IsDK | -0.316 | (-0.412, -0.223) | 0.168 | 1.183 | (0.952, 1.509) |
|  |  | LowEduc | -0.185 | (-0.361, -0.014) | 0.147 | 1.159 | (0.924, 1.477) |
|  |  | Smoke100 | -0.081 | (-0.163, 0.004) | 0.220 | 1.246 | (1.065, 1.458) |
|  |  | APOE4 | 0.053 | (-0.045, 0.158) | 0.171 | 1.186 | (0.978, 1.458) |
|  |  | MedsDiab | -0.074 | (-0.204, 0.062) |  |  |  |
|  |  | MedsHtn | -0.014 | (-0.116, 0.087) |  |  |  |
|  |  | MedsLipid | -0.250 | (-0.330, -0.168) |  |  |  |
|  |  | MedsNitro | -0.044 | (-0.153, 0.066) |  |  |  |
|  |  | PC1 |  |  | 0.064 | 1.066 | (0.848, 1.364) |
|  |  | PC2 |  |  | -0.053 | 0.948 | (0.000, 1.214) |
| intslope | Females | LPC_randomint |  |  | -0.382 | 0.682 | (0.585, 0.813) |
|  |  | LPC_randomslope |  |  | -0.574 | 0.563 | (0.000, 408.544) |
|  |  | Intercept_longit | 0.933 | (0.728, 1.145) |  |  |  |
|  |  | AgeV1 | -0.011 | (-0.014, -0.008) | 0.129 | 1.138 | (1.129, 1.150) |
|  |  | TimeV1 | 0.013 | (0.006, 0.019) |  |  |  |
|  |  | IsDK | -0.304 | (-0.396, -0.210) | 0.089 | 1.094 | (0.880, 1.318) |
|  |  | LowEduc | -0.008 | (-0.122, 0.105) | 0.117 | 1.124 | (0.924, 1.376) |
|  |  | Smoke100 | -0.067 | (-0.152, 0.010) | 0.151 | 1.163 | (1.008, 1.397) |
|  |  | APOE4 | -0.071 | (-0.171, 0.034) | 0.238 | 1.269 | (1.039, 1.612) |
|  |  | MedsDiab | -0.185 | (-0.345, -0.056) |  |  |  |
|  |  | MedsHtn | -0.035 | (-0.117, 0.034) |  |  |  |
|  |  | MedsLipid | -0.216 | (-0.293, -0.136) |  |  |  |
|  |  | MedsNitro | 0.016 | (-0.063, 0.113) |  |  |  |
|  |  | PC1 |  |  | 0.185 | 1.204 | (0.997, 1.519) |
|  |  | PC2 |  |  | 0.170 | 1.185 | (0.978, 1.450) |
|  | Males | LPC_randomint |  |  | -0.383 | 0.682 |  |
|  |  | LPC_randomslope |  |  | -0.142 | 0.868 |  |
|  |  | Intercept_longit | 1.111 |  |  |  |  |
|  |  | AgeV1 | -0.014 |  | 0.130 | 1.139 |  |
|  |  | TimeV1 | 0.011 |  |  |  |  |
|  |  | IsDK | -0.310 |  | 0.160 | 1.174 |  |
|  |  | LowEduc | -0.184 |  | 0.145 | 1.156 |  |
|  |  | Smoke100 | -0.082 |  | 0.213 | 1.237 |  |
|  |  | APOE4 | 0.050 |  | 0.176 | 1.193 |  |
|  |  | MedsDiab | -0.080 |  |  |  |  |
|  |  | MedsHtn | -0.008 |  |  |  |  |
|  |  | MedsLipid | -0.253 |  |  |  |  |
|  |  | MedsNitro | -0.050 |  |  |  |  |
|  |  | PC1 |  |  | 0.063 | 1.065 |  |
|  |  | PC2 |  |  | -0.050 | 0.951 |  |

**Notes:** **Model** – type of joint model (int – random intercept of LPC in survival sub-model; intslope – random intercept and slope of LPC in survival sub-model); **Variable** – LPC_randomint: random intercept of the metabolite (in **Survival** only), LPC_randomslope: random slope of the metabolite (in **Survival** only), Intercept_longit: intercept (in **Longitudinal** only), AgeV1: age at visit 1, TimeV1: time since visit 1, SexM: sex (1 – male, 0 – female), IsDK: country (1 – Denmark, 0 – USA), LowEduc: low education (1 – below high school, 0 – otherwise), Smoke100: smoking (1 – smoked 100 cigarettes in lifetime, 0 - otherwise), APOE4: *APOE* ɛ4 carrier status (1 – carrier, 0 – non-carrier), MedsDiab: diabetes mellitus medications (1 – taking; 0 – not taking) (in **Longitudinal** only), MedsHtn: hypertension medications (1 – taking; 0 – not taking) (in **Longitudinal** only), MedsLipid: lipid lowering medications (1 – taking; 0 – not taking) (in **Longitudinal** only), MedsNitro: angina medications (1 – taking; 0 – not taking) (in **Longitudinal** only), PC1, PC2: principal components computed from LLFS whole-genome sequencing data (in **Survival** only); **Longitudinal** – estimates for the longitudinal sub-model of JM (modeling the metabolite); **Survival** – estimates for the survival sub-model of JM (modeling mortality rate); **Beta** – estimates of regression parameters for **Variable** in respective sub-models; **HR** – hazard ratios (for a unit increase in all variables except PCs where they are for an increase by a standard deviation) computed from **Beta** (in **Survival** only); **95% CI** – respective 95% confidence intervals. The JM were estimated using R-package *joineR*. Note that confidence intervals for intslope in males were not estimated due to technical issues with the estimation procedure in *joineR*. LPC values were log-transformed and standardized (see **Data**).

**Table S6:** Results of applications of stochastic process models to measurements of LPC 15:0/0:0 and mortality data in the LLFS: Estimates of parameters in different models

| **Model** | $\ln\boldsymbol{a}_{\boldsymbol{\mu}_{\boldsymbol{0}}}$ | $\boldsymbol{b}_{\boldsymbol{\mu}_{\boldsymbol{0}}}$ | $\boldsymbol{\beta}_{\boldsymbol{\mu}_{\boldsymbol{0}}}$**(1)** | $\boldsymbol{\beta}_{\boldsymbol{\mu}_{\boldsymbol{0}}}$**(2)** | $\boldsymbol{\beta}_{\boldsymbol{\mu}_{\boldsymbol{0}}}$**(3)** | $\boldsymbol{\beta}_{\boldsymbol{\mu}_{\boldsymbol{0}}}$**(4)** | $\boldsymbol{\beta}_{\boldsymbol{\mu}_{\boldsymbol{0}}}$**(5)** | $\boldsymbol{\beta}_{\boldsymbol{\mu}_{\boldsymbol{0}}}$**(6)** | $\boldsymbol{\beta}_{\boldsymbol{\mu}_{\boldsymbol{0}}}$**(7)** | $\boldsymbol{\beta}_{\boldsymbol{\mu}_{\boldsymbol{0}}}$**(8)** | $\boldsymbol{\beta}_{\boldsymbol{\mu}_{\boldsymbol{0}}}$**(9)** | $\boldsymbol{\beta}_{\boldsymbol{\mu}_{\boldsymbol{0}}}$**(10)** | $\boldsymbol{\beta}_{\boldsymbol{\mu}_{\boldsymbol{0}}}$**(11)** |
| --- | --- | --- | --- | --- | --- | --- | --- | --- | --- | --- | --- | --- | --- |
| Unrestricted | -7.928 | 0.169 | 0.469 | 0.023 | 0.066 | 0.176 | -0.407 | -0.221 | 0.662 | 0.470 | 0.391 | -0.654 | -0.540 |
| Qzero | -7.608 | 0.136 | 0.368 | 0.055 | 0.125 | 0.178 | -0.322 | -0.146 | 0.541 | 0.384 | 0.273 | 0.111 | 0.414 |
| QnoT | -7.823 | 0.162 | 0.458 | 0.038 | 0.079 | 0.173 | -0.359 | -0.187 | 0.617 | 0.448 | 0.364 | -0.476 | -0.386 |
| QnoC | -7.912 | 0.168 | 0.482 | 0.025 | 0.067 | 0.175 | -0.404 | -0.222 | 0.663 | 0.468 | 0.388 | -0.649 | -0.543 |
| AnoT | -7.917 | 0.169 | 0.469 | 0.023 | 0.066 | 0.176 | -0.407 | -0.221 | 0.662 | 0.470 | 0.391 | -0.659 | -0.552 |
| AnoC | -7.914 | 0.169 | 0.469 | 0.023 | 0.066 | 0.176 | -0.407 | -0.221 | 0.662 | 0.470 | 0.391 | -0.661 | -0.555 |
| BnoC | -7.929 | 0.169 | 0.469 | 0.023 | 0.066 | 0.176 | -0.407 | -0.221 | 0.662 | 0.470 | 0.391 | -0.653 | -0.540 |
| F1noT | -7.918 | 0.169 | 0.469 | 0.023 | 0.066 | 0.176 | -0.407 | -0.221 | 0.662 | 0.470 | 0.391 | -0.659 | -0.550 |
| F1noC | -7.926 | 0.169 | 0.469 | 0.023 | 0.066 | 0.176 | -0.407 | -0.221 | 0.662 | 0.470 | 0.391 | -0.655 | -0.543 |
| F0noT | -7.875 | 0.166 | 0.434 | 0.025 | 0.069 | 0.171 | -0.382 | -0.197 | 0.632 | 0.463 | 0.382 | -0.587 | -0.490 |
| F0noC | -7.894 | 0.168 | 0.496 | 0.028 | 0.067 | 0.175 | -0.406 | -0.226 | 0.667 | 0.467 | 0.388 | -0.670 | -0.573 |
| ALzero | -7.705 | 0.141 | 0.405 | 0.067 | 0.115 | 0.179 | -0.319 | -0.148 | 0.551 | 0.393 | 0.291 | 0.029 | 0.296 |
| ALnoT | -7.881 | 0.166 | 0.434 | 0.025 | 0.069 | 0.171 | -0.382 | -0.197 | 0.632 | 0.463 | 0.382 | -0.584 | -0.483 |

**Table S6 (continued):**

| **Model** | $\boldsymbol{a}_{\boldsymbol{Q}}\boldsymbol{\cdot}\boldsymbol{10}^{\boldsymbol{3}}$ | $\boldsymbol{b}_{\boldsymbol{Q}}\boldsymbol{\cdot}\boldsymbol{10}^{\boldsymbol{4}}$ | $\boldsymbol{\beta}_{\boldsymbol{Q}}\boldsymbol{\cdot}\boldsymbol{10}^{\boldsymbol{3}}$ | $\boldsymbol{a}_{\boldsymbol{Y}}$ | $\boldsymbol{b}_{\boldsymbol{Y}}$  $\boldsymbol{\cdot}\boldsymbol{10}^{\boldsymbol{8}}$ | $\boldsymbol{\beta}_{\boldsymbol{Y}}$ | $\boldsymbol{\sigma}_{\boldsymbol{0}}$ | $\boldsymbol{\sigma}_{\boldsymbol{1}}$ | $\boldsymbol{\beta}_{\boldsymbol{W}}$ | $\boldsymbol{a}_{\boldsymbol{f}\boldsymbol{1}}$ | $\boldsymbol{b}_{\boldsymbol{f}\boldsymbol{1}}$ | $\boldsymbol{\beta}_{\boldsymbol{f}\boldsymbol{1}}$ | $\boldsymbol{a}_{\boldsymbol{f}\boldsymbol{0}}$ | $\boldsymbol{b}_{\boldsymbol{f}\boldsymbol{0}}$ | $\boldsymbol{\beta}_{\boldsymbol{f}\boldsymbol{0}}$ | **LogLik** |
| --- | --- | --- | --- | --- | --- | --- | --- | --- | --- | --- | --- | --- | --- | --- | --- | --- |
| Unrestricted | -2.016 | 0.578 | -0.585 | -0.053 | 0.387 | -0.025 | 0.985 | 0.270 | 0.049 | 0.284 | -0.014 | -0.107 | 1.320 | 0.023 | 0.395 | -9,964.33 |
| Qzero |  |  |  | -0.053 | 1.933 | -0.025 | 0.985 | 0.270 | 0.049 | 0.284 | -0.014 | -0.107 | -1.723 | -0.081 | -0.196 | -10,014.53 |
| QnoT | 1.324 |  | -0.031 | -0.053 | 0.077 | -0.025 | 0.985 | 0.270 | 0.049 | 0.284 | -0.014 | -0.107 | 1.205 | 0.032 | -0.069 | -9,970.63 |
| QnoC | -1.702 | 0.495 |  | -0.053 | 0.387 | -0.025 | 0.985 | 0.270 | 0.049 | 0.284 | -0.014 | -0.107 | 1.329 | 0.030 | -0.047 | -9,965.01 |
| AnoT | -2.017 | 0.578 | -0.585 | -0.053 |  | -0.025 | 0.985 | 0.270 | 0.049 | 0.284 | -0.014 | -0.107 | 1.320 | 0.023 | 0.396 | -9,964.33 |
| AnoC | -2.017 | 0.578 | -0.585 | -0.062 | 0.075 |  | 0.985 | 0.271 | 0.051 | 0.289 | -0.014 | -0.121 | 1.320 | 0.023 | 0.396 | -9,976.99 |
| BnoC | -2.017 | 0.578 | -0.585 | -0.053 | 0.111 | -0.025 | 0.984 | 0.291 |  | 0.279 | -0.014 | -0.100 | 1.320 | 0.023 | 0.396 | -9,976.35 |
| F1noT | -2.017 | 0.578 | -0.585 | -0.050 | 0.416 | -0.025 | 1.004 | 0.273 | 0.048 | 0.013 |  | -0.124 | 1.320 | 0.023 | 0.395 | -10,062.28 |
| F1noC | -2.017 | 0.578 | -0.585 | -0.052 | 0.096 | -0.027 | 0.986 | 0.270 | 0.047 | 0.237 | -0.014 |  | 1.320 | 0.023 | 0.396 | -9,971.06 |
| F0noT | -2.103 | 0.585 | -0.709 | -0.053 | 0.097 | -0.025 | 0.985 | 0.270 | 0.049 | 0.284 | -0.014 | -0.107 | 1.598 |  | 0.671 | -9,966.28 |
| F0noC | -1.803 | 0.524 | -0.235 | -0.053 | 0.097 | -0.025 | 0.985 | 0.270 | 0.049 | 0.284 | -0.014 | -0.107 | 1.332 | 0.029 |  | -9,964.67 |
| ALzero | 1.338 | -0.056 | -0.727 | -0.053 | 0.019 | -0.025 | 0.985 | 0.269 | 0.049 | 0.293 | -0.014 | -0.111 |  |  |  | -10,004.10 |
| ALnoT | -2.103 | 0.585 | -0.709 | -0.050 | 0.083 | -0.025 | 1.004 | 0.273 | 0.048 | 0.013 |  | -0.124 | 1.598 |  | 0.671 | -10,064.23 |

**Notes:** Unrestricted – main model with no restrictions on parameters; other models contain one or more restrictions on parameters to test respective null hypotheses (H0’s): H0: $Q\left( t, c \right)=0$ (Qzero); H0: $Q\left( t, c \right)=Q\left( c \right)$ (QnoT); H0: $Q\left( t, c \right)=Q\left( t \right)$ (QnoC); H0: $a\left( t, c \right)=a\left( c \right)$ (AnoT); H0: $a\left( t, c \right)=a\left( t \right)$ (AnoC); H0: $b\left( t, c \right)=b\left( t \right)$ (BnoC); H0: $f_{1}\left( t, c \right)=f_{1}\left( c \right)$ (F1noT); H0: $f_{1}\left( t, c \right)=f_{1}\left( t \right)$ (F1noC); H0: $f_{0}\left( t, c \right)=f_{0}\left( c \right)$ (F0noT); H0: $f_{0}\left( t, c \right)=f_{0}\left( t \right)$ (F0noC); H0: $f_{1}\left( t, c \right)=f_{0}\left( t,c \right)$, i.e., $AL\left( t, c \right)=0$ (ALzero); H0: $f_{1}\left( t, c \right)=f_{1}\left( c \right)$ and $f_{0}\left( t, c \right)=f_{0}\left( c \right)$, i.e., $AL\left( t, c \right)=AL\left( c \right)$ (ALnoT). Columns $\boldsymbol{\beta}_{\boldsymbol{\mu}_{\boldsymbol{0}}}\boldsymbol{(.)}$ show coefficients for the variables in the hazard rate, in the following order: (1) SexM: sex (1 – male, 0 – female); (2) IsDK: country (1 – Denmark, 0 – USA); (3) LowEduc: low education (1 – below high school, 0 – otherwise); (4) Smoke100: smoking (1 – smoked 100 cigarettes in lifetime, 0 - otherwise); (5) MedsLipid: lipid lowering medications (1 – taking; 0 – not taking); (6) MedsHtn: hypertension medications (1 – taking; 0 – not taking); (7) MedsNitro: angina medications (1 – taking; 0 – not taking); (8) MedsDiab: diabetes mellitus medications (1 – taking; 0 – not taking); (9) APOE4: *APOE* ɛ4 carrier status (1 – carrier, 0 – non-carrier); (10)-(11) PC1-PC2: first two principal components computed from LLFS whole-genome sequencing data. In other components, respective $\beta's$ show coefficients for variable SexM. The estimates of some parameters are rescaled for better visibility, see names of the columns. **LogLik:** logarithm of the likelihood function. LPC values were log-transformed and standardized (see **Data**).

**Table S7:** Applications of joint models to measurements of LPC variants and mortality data in the LLFS: Estimates from familial bootstrap

| **Metabolite** | **Median HR [Range]** | | |
| --- | --- | --- | --- |
|  | **Total** | **Females** | **Males** |
| LPC 0:0/16:0 | **0.831 [0.674, 0.936]** | **0.833 [0.672, 0.956]** | **0.840 [0.583, 0.946]** |
| LPC 0:0/16:1 | 0.980 [0.831, 1.203] | 0.961 [0.693, 1.243] | 0.978 [0.786, 1.299] |
| LPC 0:0/18:0 | 0.960 [0.834, 1.128] | 0.967 [0.783, 1.157] | 0.935 [0.786, 1.148] |
| LPC 0:0/18:1 | 0.866 [0.743, 1.013] | 0.896 [0.755, 1.047] | 0.854 [0.679, 1.042] |
| LPC 0:0/18:2 | **0.792 [0.669, 0.932]** | 0.853 [0.655, 1.003] | **0.728 [0.578, 0.851]** |
| LPC 0:0/20:3 | **0.860 [0.731, 0.995]** | 0.842 [0.678, 1.046] | 0.854 [0.566, 1.057] |
| LPC 0:0/20:4 | **0.826 [0.739, 0.948]** | **0.782 [0.592, 0.934]** | **0.866 [0.727, 0.990]** |
| LPC 0:0/22:6 | **0.795 [0.706, 0.900]** | **0.737 [0.603, 0.894]** | 0.860 [0.707, 1.040] |
| LPC 14:0/0:0 | **0.819 [0.692, 0.950]** | 0.839 [0.628, 1.004] | **0.812 [0.532, 0.960]** |
| LPC 15:0/0:0 | **0.743 [0.585, 0.849]** | **0.749 [0.609, 0.869]** | **0.763 [0.537, 0.887]** |
| LPC 16:0/0:0 | **0.809 [0.605, 0.924]** | 0.831 [0.621, 1.035] | **0.811 [0.468, 0.934]** |
| LPC 16:1/0:0 | 0.956 [0.822, 1.076] | 0.956 [0.786, 1.104] | 0.954 [0.788, 1.152] |
| LPC 17:0/0:0 | **0.816 [0.689, 0.961]** | **0.837 [0.634, 0.963]** | **0.786 [0.596, 0.953]** |
| LPC 18:0/0:0 | 1.014 [0.891, 1.145] | 1.010 [0.858, 1.204] | 1.000 [0.849, 1.186] |
| LPC 18:1/0:0 | **0.830 [0.623, 0.931]** | **0.835 [0.642, 0.976]** | **0.834 [0.564, 0.946]** |
| LPC 18:2/0:0 | **0.815 [0.669, 0.940]** | 0.856 [0.612, 1.080] | **0.795 [0.362, 0.951]** |
| LPC 18:3/0:0 | 0.860 [0.744, 1.019] | 0.966 [0.750, 1.309] | **0.734 [0.254, 0.960]** |
| LPC 20:2/0:0 | **0.803 [0.690, 0.935]** | 0.840 [0.701, 1.000] | **0.785 [0.585, 0.944]** |
| LPC 20:3/0:0 | **0.772 [0.661, 0.871]** | **0.776 [0.645, 0.925]** | **0.769 [0.540, 0.931]** |
| LPC 20:4/0:0 | **0.819 [0.733, 0.922]** | **0.808 [0.598, 0.977]** | **0.842 [0.649, 0.967]** |
| LPC 20:5/0:0 | **0.738 [0.658, 0.832]** | **0.704 [0.588, 0.874]** | **0.783 [0.582, 0.903]** |
| LPC 22:5/0:0 | **0.797 [0.715, 0.900]** | **0.764 [0.618, 0.906]** | **0.820 [0.654, 0.936]** |
| LPC 22:6/0:0 | **0.771 [0.695, 0.847]** | **0.733 [0.618, 0.861]** | **0.806 [0.707, 0.979]** |

**Notes:** The table reports medians of hazard ratios (HR) for a unit increase in the log-transformed and standardized metabolite values (ranges in the parentheses) for the association parameters for the metabolites in the survival sub-model computed in using the familial bootstrap method [1] in 100 bootstrap samples generated from the original sample. See main text (section **MATERIALS AND METHODS:** **Sensitivity analyses**) for details. The joint models were estimated using R-package *JM*. The cases where the range of HR does not contain one are highlighted in **bold**. The cases where 95% confidence intervals (CI) for HR in the main calculations (Table 2) did not contain one but the HR range in the familial bootstrap included one are highlighted in yellow background. The case where 95% CI for HR in the main calculations (Table 2) contained one but the HR range in the familial bootstrap did not include one is highlighted in grey background.

**Table S8:** Applications of stochastic process models to measurements of LPC 15:0/0:0 and mortality data in the LLFS: Estimates from familial bootstrap

| **Model** | $\ln\boldsymbol{a}_{\boldsymbol{\mu}_{\boldsymbol{0}}}$ | $\boldsymbol{b}_{\boldsymbol{\mu}_{\boldsymbol{0}}}$ | $\boldsymbol{\beta}_{\boldsymbol{\mu}_{\boldsymbol{0}}}$**(1)** | $\boldsymbol{\beta}_{\boldsymbol{\mu}_{\boldsymbol{0}}}$**(2)** | $\boldsymbol{\beta}_{\boldsymbol{\mu}_{\boldsymbol{0}}}$**(3)** | $\boldsymbol{\beta}_{\boldsymbol{\mu}_{\boldsymbol{0}}}$**(4)** | $\boldsymbol{\beta}_{\boldsymbol{\mu}_{\boldsymbol{0}}}$**(5)** | $\boldsymbol{\beta}_{\boldsymbol{\mu}_{\boldsymbol{0}}}$**(6)** | $\boldsymbol{\beta}_{\boldsymbol{\mu}_{\boldsymbol{0}}}$**(7)** | $\boldsymbol{\beta}_{\boldsymbol{\mu}_{\boldsymbol{0}}}$**(8)** | $\boldsymbol{\beta}_{\boldsymbol{\mu}_{\boldsymbol{0}}}$**(9)** | $\boldsymbol{\beta}_{\boldsymbol{\mu}_{\boldsymbol{0}}}$**(10)** | $\boldsymbol{\beta}_{\boldsymbol{\mu}_{\boldsymbol{0}}}$**(11)** |
| --- | --- | --- | --- | --- | --- | --- | --- | --- | --- | --- | --- | --- | --- |
| Unrestricted | -8.778 (3.363) | 0.172 (0.008) | 0.474 (0.118) | 0.021 (0.163) | 0.081 (0.139) | 0.186 (0.109) | -0.408 (0.121) | -0.231 (0.112) | 0.690 (0.123) | 0.481 (0.205) | 0.394 (0.132) | -0.169 (1.869) | -0.060 (3.763) |
| Qzero | -8.060 (2.609) | 0.136 (0.004) | 0.363 (0.072) | 0.051 (0.100) | 0.137 (0.110) | 0.184 (0.080) | -0.315 (0.075) | -0.148 (0.083) | 0.549 (0.086) | 0.383 (0.138) | 0.258 (0.089) | 0.445 (1.432) | 0.664 (2.840) |
| QnoT | -8.487 (3.074) | 0.164 (0.007) | 0.461 (0.102) | 0.037 (0.141) | 0.091 (0.130) | 0.180 (0.098) | -0.354 (0.102) | -0.192 (0.100) | 0.637 (0.108) | 0.454 (0.182) | 0.363 (0.118) | -0.072 (1.697) | -0.049 (3.422) |
| QnoC | -8.744 (3.349) | 0.171 (0.008) | 0.488 (0.113) | 0.023 (0.160) | 0.082 (0.139) | 0.185 (0.108) | -0.405 (0.121) | -0.231 (0.112) | 0.690 (0.122) | 0.476 (0.204) | 0.390 (0.131) | -0.167 (1.861) | -0.066 (3.754) |
| AnoT | -8.777 (3.348) | 0.172 (0.008) | 0.473 (0.118) | 0.020 (0.163) | 0.081 (0.139) | 0.186 (0.108) | -0.408 (0.121) | -0.231 (0.112) | 0.690 (0.123) | 0.480 (0.205) | 0.394 (0.132) | -0.167 (1.853) | -0.057 (3.732) |
| AnoC | -8.774 (3.361) | 0.172 (0.008) | 0.473 (0.118) | 0.020 (0.163) | 0.081 (0.139) | 0.186 (0.108) | -0.408 (0.121) | -0.231 (0.112) | 0.690 (0.123) | 0.480 (0.205) | 0.394 (0.132) | -0.167 (1.861) | -0.058 (3.747) |
| BnoC | -8.781 (3.345) | 0.172 (0.008) | 0.472 (0.117) | 0.019 (0.162) | 0.081 (0.139) | 0.186 (0.108) | -0.410 (0.121) | -0.231 (0.112) | 0.690 (0.123) | 0.480 (0.205) | 0.394 (0.132) | -0.160 (1.857) | -0.046 (3.740) |
| F1noT | -8.769 (3.369) | 0.172 (0.009) | 0.475 (0.116) | 0.021 (0.163) | 0.081 (0.140) | 0.186 (0.109) | -0.407 (0.121) | -0.230 (0.112) | 0.689 (0.123) | 0.480 (0.206) | 0.392 (0.131) | -0.166 (1.867) | -0.058 (3.762) |
| F1noC | -8.780 (3.350) | 0.172 (0.009) | 0.473 (0.118) | 0.020 (0.163) | 0.081 (0.139) | 0.186 (0.109) | -0.409 (0.120) | -0.231 (0.112) | 0.691 (0.123) | 0.480 (0.205) | 0.394 (0.132) | -0.164 (1.863) | -0.049 (3.745) |
| F0noT | -8.630 (3.198) | 0.170 (0.008) | 0.441 (0.109) | 0.026 (0.151) | 0.082 (0.134) | 0.179 (0.103) | -0.377 (0.110) | -0.203 (0.105) | 0.654 (0.112) | 0.472 (0.193) | 0.383 (0.124) | -0.155 (1.771) | -0.093 (3.558) |
| F0noC | -8.725 (3.351) | 0.171 (0.008) | 0.501 (0.110) | 0.025 (0.162) | 0.082 (0.140) | 0.186 (0.109) | -0.407 (0.122) | -0.235 (0.112) | 0.694 (0.122) | 0.475 (0.205) | 0.389 (0.132) | -0.187 (1.860) | -0.093 (3.749) |
| ALzero | -8.202 (2.706) | 0.142 (0.005) | 0.400 (0.076) | 0.066 (0.106) | 0.126 (0.114) | 0.184 (0.083) | -0.311 (0.079) | -0.151 (0.085) | 0.561 (0.089) | 0.396 (0.145) | 0.282 (0.094) | 0.367 (1.482) | 0.555 (2.957) |
| ALnoT | -8.633 (3.192) | 0.170 (0.008) | 0.441 (0.110) | 0.026 (0.151) | 0.082 (0.134) | 0.179 (0.103) | -0.377 (0.110) | -0.203 (0.105) | 0.654 (0.112) | 0.472 (0.193) | 0.383 (0.124) | -0.154 (1.769) | -0.090 (3.550) |

**Table S8 (continued):**

| **Model** | $\boldsymbol{a}_{\boldsymbol{Q}}\boldsymbol{\cdot}\boldsymbol{10}^{\boldsymbol{3}}$ | $\boldsymbol{b}_{\boldsymbol{Q}}\boldsymbol{\cdot}\boldsymbol{10}^{\boldsymbol{4}}$ | $\boldsymbol{\beta}_{\boldsymbol{Q}}\boldsymbol{\cdot}\boldsymbol{10}^{\boldsymbol{3}}$ | $\boldsymbol{a}_{\boldsymbol{Y}}$ | $\boldsymbol{b}_{\boldsymbol{Y}}$  $\boldsymbol{\cdot}\boldsymbol{10}^{\boldsymbol{8}}$ | $\boldsymbol{\beta}_{\boldsymbol{Y}}$ | $\boldsymbol{\sigma}_{\boldsymbol{0}}$ | $\boldsymbol{\sigma}_{\boldsymbol{1}}$ | $\boldsymbol{\beta}_{\boldsymbol{W}}$ | $\boldsymbol{a}_{\boldsymbol{f}\boldsymbol{1}}$ | $\boldsymbol{b}_{\boldsymbol{f}\boldsymbol{1}}$ | $\boldsymbol{\beta}_{\boldsymbol{f}\boldsymbol{1}}$ | $\boldsymbol{a}_{\boldsymbol{f}\boldsymbol{0}}$ | $\boldsymbol{b}_{\boldsymbol{f}\boldsymbol{0}}$ | $\boldsymbol{\beta}_{\boldsymbol{f}\boldsymbol{0}}$ |
| --- | --- | --- | --- | --- | --- | --- | --- | --- | --- | --- | --- | --- | --- | --- | --- |
| Unrestricted | -2.311 (1.188) | 0.656 (0.259) | -0.001 (0.000) | -0.053 (0.005) | 0.008 (0.037) | -0.022 (0.010) | 0.983 (0.024) | 0.269 (0.008) | 0.038 (0.031) | 0.282 (0.039) | -0.014 (0.001) | -0.106 (0.029) | 1.284 (0.382) | 0.023 (0.008) | 0.359 (0.368) |
| Qzero |  |  |  | -0.053 (0.005) | 0.008 (0.037) | -0.022 (0.010) | 0.983 (0.024) | 0.269 (0.008) | 0.038 (0.031) | 0.282 (0.039) | -0.014 (0.001) | -0.106 (0.029) | -1.294 (0.798) | -0.089 (0.013) | -0.136 (0.086) |
| QnoT | 1.474 (0.436) |  | 0.000 (0.001) | -0.053 (0.005) | 0.008 (0.037) | -0.022 (0.010) | 0.983 (0.024) | 0.269 (0.008) | 0.038 (0.031) | 0.282 (0.039) | -0.014 (0.001) | -0.106 (0.029) | 1.197 (0.324) | 0.031 (0.006) | -0.131 (0.289) |
| QnoC | -2.088 (1.179) | 0.586 (0.247) |  | -0.053 (0.005) | 0.008 (0.037) | -0.022 (0.010) | 0.983 (0.024) | 0.269 (0.008) | 0.038 (0.031) | 0.282 (0.039) | -0.014 (0.001) | -0.106 (0.029) | 1.303 (0.374) | 0.028 (0.007) | -0.074 (0.237) |
| AnoT | -2.288 (1.169) | 0.655 (0.253) | -0.001 (0.000) | -0.053 (0.004) |  | -0.022 (0.010) | 0.983 (0.024) | 0.269 (0.008) | 0.038 (0.031) | 0.282 (0.039) | -0.014 (0.001) | -0.106 (0.029) | 1.272 (0.395) | 0.024 (0.009) | 0.373 (0.363) |
| AnoC | -2.298 (1.171) | 0.658 (0.254) | -0.001 (0.000) | -0.062 (0.005) | 0.005 (0.030) |  | 0.983 (0.024) | 0.270 (0.008) | 0.040 (0.032) | 0.287 (0.040) | -0.014 (0.001) | -0.119 (0.029) | 1.266 (0.401) | 0.024 (0.009) | 0.367 (0.356) |
| BnoC | -2.354 (1.235) | 0.671 (0.271) | -0.001 (0.000) | -0.053 (0.005) | 0.010 (0.049) | -0.023 (0.010) | 0.983 (0.024) | 0.286 (0.014) |  | 0.279 (0.040) | -0.014 (0.001) | -0.101 (0.029) | 1.241 (0.428) | 0.024 (0.009) | 0.362 (0.356) |
| F1noT | -2.256 (1.199) | 0.647 (0.258) | -0.001 (0.000) | -0.050 (0.004) | 0.005 (0.033) | -0.022 (0.010) | 1.002 (0.024) | 0.273 (0.008) | 0.037 (0.030) | 0.013 (0.032) |  | -0.123 (0.030) | 1.278 (0.386) | 0.023 (0.009) | 0.327 (0.458) |
| F1noC | -2.346 (1.257) | 0.669 (0.267) | -0.001 (0.001) | -0.052 (0.005) | 0.008 (0.037) | -0.024 (0.010) | 0.984 (0.024) | 0.270 (0.008) | 0.036 (0.031) | 0.236 (0.038) | -0.014 (0.001) |  | 1.245 (0.418) | 0.024 (0.009) | 0.358 (0.359) |
| F0noT | -2.279 (1.047) | 0.627 (0.220) | -0.001 (0.000) | -0.053 (0.005) | 0.008 (0.037) | -0.022 (0.010) | 0.983 (0.024) | 0.269 (0.008) | 0.038 (0.031) | 0.282 (0.039) | -0.014 (0.001) | -0.106 (0.029) | 1.650 (0.306) |  | 0.586 (0.414) |
| F0noC | -2.140 (1.093) | 0.610 (0.231) | -0.000 (0.000) | -0.053 (0.005) | 0.008 (0.037) | -0.022 (0.010) | 0.983 (0.024) | 0.269 (0.008) | 0.038 (0.031) | 0.282 (0.039) | -0.014 (0.001) | -0.106 (0.029) | 1.282 (0.370) | 0.029 (0.008) |  |
| ALzero | 1.574 (0.810) | -0.080 (0.096) | -0.001 (0.001) | -0.053 (0.005) | 0.008 (0.037) | -0.022 (0.010) | 0.983 (0.024) | 0.269 (0.008) | 0.038 (0.031) | 0.292 (0.039) | -0.014 (0.001) | -0.109 (0.030) |  |  |  |
| ALnoT | -2.295 (1.047) | 0.629 (0.219) | -0.001 (0.000) | -0.050 (0.004) | 0.005 (0.033) | -0.022 (0.010) | 1.002 (0.024) | 0.273 (0.008) | 0.037 (0.030) | 0.013 (0.032) |  | -0.123 (0.030) | 1.653 (0.315) |  | 0.579 (0.428) |

**Notes:** The table reports means and standard deviations (in parentheses) of respective parameter estimates computed in using the familial bootstrap method [1] in 100 bootstrap samples generated from the original sample. See main text (section **MATERIALS AND METHODS:** **Sensitivity analyses**) for details. See explanations of the models and parameters in Notes under **Table S6**.

**References:**

1.	Borecki IB and Province MA. Genetic and Genomic Discovery Using Family Studies. Circulation. 2008; 118(10):1057-1063.

### Code implementing likelihood estimation procedure of SPM

This document describes the code that can be used to estimate the likelihood function of Stochastic Process Model (SPM) used in the paper. It provides the code for "unrestricted" model that can be modified to specify one or more restrictions on parameters to perform hypotheses testing presented in the text.

The function estimates the likelihood for the following SPM:

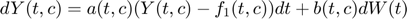

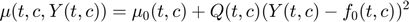

with the following specification of components:

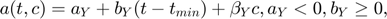

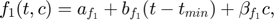

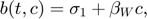

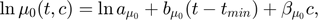

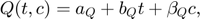

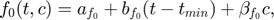

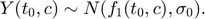

#### Syntax:

function lnLik = LogLik(Param, DataSPM, t_min, NamesCovar)

**Parameters:**

Param – a column vector of model parameters in the following order:

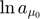
,
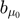
,
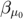
,
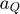
,
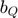
,
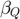
,
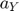
,
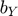
,
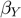
,
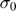
,
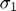
,
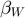
,
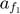
,
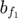
,
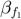
,
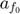
,
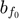
,
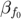

DataSPM – a table with the following variables (in any order; can have additional variables which will be ignored): *Age* (start of age interval), *AgeNext* (end of age interval), *IndicatorEvent* (a binary variable indicating event [1] or no event [0] within the age interval (*Age* , *AgeNext*)), *Yt* (longitudinal variable modelled by *Y*(*t*, *c*)), *IsFirstRow* (a binary variable indicating the first record for an individual: 1 – first record; 0 – otherwise), *IsLastRow* (a binary variable indicating the last record for an individual: 1 – last record; 0 – otherwise), and variables to be included as additiona covariates (*c*), see NamesCovar

t_min – minimal age used in formulas, see above

NamesCovar – cell array with names of variables in DataSPM to be used as additional covariates (*c*): the first cell contains names of variables to be used as covariates in
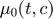
 and the second cell contains names of variables to be used as covariates in other components

**Output:**

lnLik – minus logarithm of the likelihood function

```
function lnLik = LogLikSPM(Param, DataSPM, t_min, NamesCovar)

NumRows=height(DataSPM);

NamesCovarMu0=NamesCovar{1};
NamesCovarOther=NamesCovar{2};
NumCovarMu0=length(NamesCovarMu0);
NumCovarOther=length(NamesCovarOther);

ln_a_mu0=Param(1);
b_mu0=Param(2);
b_covar_mu0=Param(3:(3+NumCovarMu0-1));
a_mu11=Param(3+NumCovarMu0);
b_mu11=Param(4+NumCovarMu0);
a_mu12=a_mu11;
b_mu12=b_mu11;
b_covar_Q=Param((5+NumCovarMu0):(5+NumCovarMu0+NumCovarOther-1));
a_Y=Param(5+NumCovarMu0+NumCovarOther);
b_Y=Param(6+NumCovarMu0+NumCovarOther);
b_covar_Y=Param((7+NumCovarMu0+NumCovarOther):(7+NumCovarMu0+2*NumCovarOther-1));
sigma0=Param(7+NumCovarMu0+2*NumCovarOther);
sigma1=Param(8+NumCovarMu0+2*NumCovarOther);
b_covar_W=Param((9+NumCovarMu0+2*NumCovarOther):(9+NumCovarMu0+3*NumCovarOther-1));
a_f1=Param(9+NumCovarMu0+3*NumCovarOther);
b_f1=Param(10+NumCovarMu0+3*NumCovarOther);
b_covar_f1=Param((11+NumCovarMu0+3*NumCovarOther):(11+NumCovarMu0+4*NumCovarOther-1));
a_f0=Param(11+NumCovarMu0+4*NumCovarOther);
b_f0=Param(12+NumCovarMu0+4*NumCovarOther);
b_covar_f0=Param((13+NumCovarMu0+4*NumCovarOther):(13+NumCovarMu0+5*NumCovarOther-1));

delta_i_all=DataSPM.IndicatorDeath;
t=DataSPM.Age;
t_next=DataSPM.AgeNext;
Yt=DataSPM.Yt;
IsFirstRow=DataSPM.IsFirstRow;
IsLastRow=DataSPM.IsLastRow;

if NumCovarOther==1
    Xt_other=DataSPM.(NamesCovarOther{1});
else
    Xt_other=NaN*ones(NumRows,NumCovarOther);
    for i=1:NumCovarOther
        Xt_other(:,i)=DataSPM.(NamesCovarOther{i});
    end
end

if NumCovarMu0==1
    Xt_mu0=DataSPM.(NamesCovarMu0{1});
else
    Xt_mu0=NaN*ones(NumRows,NumCovarMu0);
    for i=1:NumCovarMu0
        Xt_mu0(:,i)=DataSPM.(NamesCovarMu0{i});
    end
end

lnLik=0;
for i=1:NumRows
    delta_i=delta_i_all(i);
    tk=t(i);
    tk_next=t_next(i);
    Ytk=Yt(i);
    Xtk_other=Xt_other(i,:);
    Xtk_mu0=Xt_mu0(i,:);

    mu0_tk=exp(ln_a_mu0+b_mu0*(tk-t_min)+Xtk_mu0*b_covar_mu0);
    f0_tk=a_f0+b_f0*(tk-t_min)+Xtk_other*b_covar_f0;
    mu11_tk=a_mu11+b_mu11*tk+Xtk_other*b_covar_Q;
    mu12_tk=a_mu12+b_mu12*tk+Xtk_other*b_covar_Q;
    if Ytk<=f0_tk
        mu1_tk=mu11_tk;
    else
        mu1_tk=mu12_tk;
    end
    mu_tk=mu0_tk+mu1_tk*(Ytk-f0_tk)^2;

    if IsFirstRow(i)==1
        lnLY=0;
        lnLQ=0;

        t0=t(i);
        Yt0=Yt(i);

        Ybar_tk_prev=a_f1+b_f1*(t0-t_min)+Xtk_other*b_covar_f1;
        if (sigma0>0)
            lnLY=lnLY-log(sqrt(2*pi)*sigma0)-((Yt0-Ybar_tk_prev)^2)/(2*sigma0^2);
        end
    else
        tk_prev=t(i-1);
        Ytk_prev=Yt(i-1);

        a_tk_prev=a_Y+b_Y*(tk_prev-t_min)+Xtk_other*b_covar_Y;
        f1_tk_prev=a_f1+b_f1*(tk_prev-t_min)+Xtk_other*b_covar_f1;

        Ybar_tk_prev=Ytk_prev+a_tk_prev*(Ytk_prev-f1_tk_prev)*(tk-tk_prev);
        sigma1_tk=sigma1+Xtk_other*b_covar_W;
        if (sigma1_tk>0) && ((tk-tk_prev)>0)
            lnLY=lnLY-log(sqrt(2*pi*(tk-tk_prev))*sigma1_tk)-((Ytk-Ybar_tk_prev)^2)/(2*(tk-tk_prev)*sigma1_tk^2);
        end
    end

    if IsLastRow(i)==1
        if delta_i==0
            lnLQ=lnLQ-mu_tk*(tk_next-tk);
        elseif delta_i==1
            if (mu_tk<=0) || (tk_next==tk)
                ln_mu_tk=log(1e-323);
            else
                ln_mu_tk=log(1-exp(-mu_tk*(tk_next-tk)));
            end
            lnLQ=lnLQ+ln_mu_tk;
        end
    else
        lnLQ=lnLQ-mu_tk*(tk_next-tk);
    end

    if IsLastRow(i)==1
        lnLik=lnLik+lnLY+lnLQ;
    end
end
lnLik=-lnLik;
```

[*Published with MATLAB® R2023b*](https://www.mathworks.com/products/matlab)
